# Vocal markers of schizophrenia: assessing the generalizability of machine learning models and their clinical applicability

**DOI:** 10.1101/2024.11.06.24316839

**Authors:** Alberto Parola, Emil Trenckner Jessen, Astrid Rybner, Marie Damsgaard Mortensen, Stine Nyhus Larsen, Arndis Simonsen, Jessica Mary Lin, Yuan Zhou, Huiling Wang, Katja Koelkebeck, Konstantinos Sechidis, Vibeke Bliksted, Riccardo Fusaroli

## Abstract

**Background and Hypothesis:** Machine Learning (ML) models have been argued to reliably predict diagnosis and symptoms of schizophrenia based on voice data only. However, it is unclear to what extent such ML markers would generalize to different clinical samples and different languages, a crucial assessment to move towards clinical applicability. In this study, we systematically assessed the generalizability of ML models of vocal markers of schizophrenia across contexts and languages.

**Study Design:** We trained models relying on a large cross-linguistic dataset (Danish, German, Chinese) of 217 patients with schizophrenia and 221 controls, and used a conservative pipeline to minimize overfitting. We tested the models’ generalizability on: (i) new participants, speaking the same language; (ii) new participants, speaking a different language; (iii) further, we assessed whether training on data with multiple languages would improve generalizability using Mixture of Expert (MoE) and multilingual models.

**Results:** Model performance was comparable to state-of-the-art findings (F1-score ∼ 0.75) within the same language; however, models did not generalize well - showing a substantial decrease - when tested on new languages. The performance of MoE and multilingual models was also generally low (F1-score ∼ 0.50).

**Conclusions:** Overall, the cross-linguistic generalizability of vocal markers of schizophrenia is limited. We argue that more emphasis should be placed on collecting large open cross-linguistic datasets to systematically test the generalizability of voice-based ML models, and on identifying more precise mechanisms of how the clinical features of schizophrenia are expressed in language and voice, and how different languages vary in that expression.

## Introduction

Voice analysis is argued to be a promising candidate marker for schizophrenia (Corona Hernández et al., 2023). Indeed, schizophrenia is associated with an atypical vocal profile characterized by a distinctive tone of voice, prolonged pauses and poverty of speech (Cohen et al., 2014; Parola, Simonsen, et al., 2020a, 2023). These acoustic features are in turn related to the symptoms of the disorders, such as alogia and flat affect (Alpert et al., 2002; Cohen, Cowan, et al., 2020; Cohen, Cox, Le, et al., 2020a), which are routinely assessed by evaluating the patient’s vocal and acoustic profile. Beyond the association with symptoms, crucially, vocal behavior provides a window into the motor, social, and cognitive impairments underlying the disorder (Palaniyappan, 2021a; Parola, Brasso, et al., 2021; Parola, Gabbatore, et al., 2021; Parola, Salvini, et al., 2020; Simonsen, et al., 2021). For example, extrapyramidal motor symptoms secondary to antipsychotic medication are reflected in reduced voice quality (e.g. increased jitter and shimmer, Hitczenko et al., 2023; see also Fusaroli et al., 2023), and the social and cognitive impairments commonly reported in schizophrenia are evident in difficulties in speech fluency (e.g., increased pauses) or in controlling the voice to express emotional content (e.g., Cohen, Cowan, et al., 2020; Cohen, Cox, Masucci, et al., 2020). The voice profile of patients thus conveys a rich source of information about the biological, clinical and social aspects of the disorder, and leveraging on this information speech-based models could potentially improve the diagnosis and prognosis of schizophrenia and allow to track mental states (Tang et al., 2023; Voleti et al., 2019), symptoms (Arevian et al., 2020; Cummins et al., 2015), cognition (Stegmann et al., 2022; Tang et al., 2023) and motor disorders (de Boer et al., 2020; Hitczenko et al., 2023) over time.

Machine learning (ML) applications have already shown their potential to identify vocal and linguistic markers of schizophrenia (e.g. Cohen, Cox, et al., 2021; Cohen, Schwartz, et al., 2021; Corcoran & Cecchi, 2020; Hitczenko et al., 2021), by identifying diagnosed individuals from controls with high accuracy (Cohen, Cox, Le, et al., 2020b; De Boer et al., 2021; Espinola et al., 2020), and accurately predicting and monitoring patients’ symptoms over time (Arevian et al., 2020; Daniel et al., 2023; Narkhede et al., 2022; Tahir et al., 2019).

However, a more attentive screening of the literature reveals some important methodological limitations. For example, both meta-analyses (Parola, Simonsen, et al., 2020b) and empirical work (Parola, Lin, et al., 2023; Parola, Simonsen, et al., 2023) have shown that the generalizability of voice-markers is limited and identified some of the reasons for these results. The datasets used for training and testing ML applications in schizophrenia are generally much smaller - large samples are on the order of 100 participants - than traditional datasets used to develop ML systems, such as automatic speech recognition systems (e.g. Krishnan Parthasarathi & Strom, 2019). Furthermore, schizophrenia is a highly heterogeneous disorder, and models trained on small samples with homogeneous features, may not generalize well when applied to patients with other clinical or socio-demographic characteristics (Chekroud et al., 2024). Finally, while vocal behavior may be largely influenced by linguistic and cultural factors, such as the morphosyntactic structure of a particular language, all studies of vocal markers of schizophrenia have investigated single monolingual samples without testing the cross-linguistic generalizability of these markers (Parola, Simonsen, et al., 2023). All these limitations can have serious consequences for the development of speech markers. First, models can overfit to specific features of the data (e.g. more female patients than controls, or only a limited range of positive symptoms in the patient group) or even random noise instead of to the true signal (Voleti et al., 2020) - which is even more likely when the number of features used is large compared to the sample size - leading to good predictions in the training data, but not when generalizing to other datasets. For example, Voleti et al. (2020) found that reported accuracy in clinical speech machine learning studies declines as a function of increasing sample size, with small sample size studies yielding an overly optimistic estimate of accuracy. Second, when multivariate speech patterns detected by complex algorithms correlate with socio-demographic, ethnic or gender characteristics, models not adequately tested may contain harmful biases that lead to misclassification of specific demographic or ethnic groups (see Hauglid, 2022; Hitczenko et al., 2023). Finally, models trained on small samples and for binary classification tasks may not learn diagnosis-specific features, but develop heuristics that are applicable to multiple conditions or only apply to samples from certain matched socio-demographic groups, resulting in low specificity and thus having a limited clinical applicability. For example, Hansen et al. (2023), found that that speech-based ML models predicting binary psychiatric diagnoses (e.g. schizophrenia vs. controls) have low specificity and rely on acoustic markers of generic differences between clinical and non-clinical populations, or markers of clinical features common to multiple clinical conditions.

Precision medicine requires models which reliably and robustly predict outcomes for new, unseen patients (Chekroud et al., 2016; Steyerberg, 2019). A clear assessment of models’ generalization performance and a thorough understanding of the capability of algorithms to adapt to various recording and data collection settings, clinical subgroups, and speech tasks, is thus a fundamental prerequisite for developing clinical applications which can affect clinical practice and improve patients’ outcomes (Chekroud et al., 2024). What is crucially lacking in schizophrenia is a systematic evaluation of the generalization performance of speech-based predictive models, to assess how robust these models are and under which conditions they generalize. First, testing the generalizability of ML models may provide a more realistic assessment of the performance of speech-based predictive models and their potential to improve clinical practice. Second, this assessment could help to disentangle which features are more robust and generalize better, and which are instead more affected by specific characteristics of a sample, or by the language spoken by patients. For example, the generalizability of results may reveal whether the identified voice patterns are related to universal biological mechanisms underlying schizophrenia and its symptoms (e.g., disorders in vocal motor control and psychomotor slowing) across contexts and languages, or whether they are instead related to specific characteristics of the speech task, experimental context and recording setting. Indeed, the identification of clinically relevant features that are less likely to overfit is a crucial step towards developing clinical applications (e.g. Voleti et al., 2020).

Third, an assessment of generalizability can identify which kind of bias may emerge when training and testing predictive models on different samples. For example, a model trained on a particular sample might be particularly poor at identifying women with schizophrenia, or participants with specific socio demographic features (e.g. Achenie et al., 2019), when tested on a sample with new participants. Finally, this assessment could also identify which methodological approaches are more robust and promising, and whether combining models from different samples (e.g. ensemble or mixture of experts (MoE) model) can make them more robust and apt to find features that are more related to the mechanisms of the disorder. Indeed, previous studies have shown that MoE ensemble models (Sechidis et al., 2021), which combine predictions of models trained on different corpora, or multi language models (Hollenstein et al., 2021) trained on multilingual corpora, can outperform models trained on single corpora and improve models’ generalization performance.

### The current study

This study specifically aims to put clinical applicability first and to investigate the generalizability of ML models of vocal markers of schizophrenia. We reviewed the current literature to identify previous studies using ML models to predict diagnosis or symptoms of schizophrenia and summarized their state-of-the art performance. We then relied on a large cross-linguistic dataset with multiple audio-recordings of patients with schizophrenia and controls in three different languages (Danish, German, Chinese) to provide a more solid basis for estimating the generalizability of voice-based ML models. The speech samples were collected using the same standardized task, i.e. the Animated Triangle Task (ATT), in the different corpora to ensure comparability. Finally, we developed a rigorous ML pipeline to minimize overfitting, including cross-validated training sets, multilanguage and MoE models, and to test the generalizability of speech-based ML models when predicting diagnosis of schizophrenia. This setup allowed us to investigate the following four main questions.

### Research questions

**Q1:** How well do voice-based ML models generalize to different participants from the same study, performing the same task and speaking the same language?

**Q2:** How well do voice-based ML models generalize to participants from a different study speaking a different native language?

**Q3:** How well does combining predictions from models trained on different languages help improve performance when assessing participants from a different study speaking a different native language?

**Q4:** To what extent does training models on a multilingual training set, i.e. combining participants from different studies speaking a different native language, help to improve performance when assessing participants from a different study speaking a different native language?

## Methods

### Participants

We collected a Danish (DK), German (GE), Chinese (CH) cross-linguistic dataset involving 217 participants with schizophrenia (105 DK, 61GE, 51CH) and 221 matched controls (HC) (116DK, 62GE, 43CH). The samples for the present study were collected in separate studies assessing implicit mentalizing (theory of mind) ability in patients with schizophrenia and healthy controls. Information on demographics, IQ, psychopathology, and social functioning is summarized in **Table 1**. Detailed information on each study is reported in the **Supplementary Material 1** (**S1)**. Note that the samples are comparable in terms of demographic and clinical features (see Table 1 and S1).

**Table 1.** Demographic and clinical characteristics of patients with schizophrenia and healthy controls (HC).

| Corpus | Danish |  | German |  | Chinese |  |
| --- | --- | --- | --- | --- | --- | --- |
| Diagnosis | SCZ<br>N = 105 | HC<br>N = 116 | SCZ<br>N = 61 | HC<br>N = 62 | SCZ<br>N = 51 | HC<br>N = 43 |
| Age | 26.51<br>(8.82) | 26.44<br>(8.96) | 31.71<br>(9.92) | 33.22<br>(8.79) | 27.22<br>(7.25) | 29.69<br>(8.72) |
| Education | 12.9<br>(2.73) | 14.86<br>(2.61) | 12.1<br>(1.48) | 12.3<br>(1.11) | 12.7<br>(2.69) | 14.12<br>(2.37) |
| Reported biological sex<br>(n. of females and %) | 45<br>(43%) | 50<br>(43%) | 22<br>(36%) | 24<br>(38%) | 23<br>(45%) | 19<br>(44%) |
| Verbal IQ | 89.13<br>(18.7) | 102.12<br>(16.1) | 112.0<br>(15.6) | 116.5<br>(16.9) | 96.3<br>(16.6) | 100.27<br>(14.7) |
| SANS total | 9.66<br>(4.41) | NA | 7.65<br>(2.79)<sup>1</sup> | NA | 7.61<br>(3.01) | NA |
| SAPS total | 10.32 (4.91) | NA | 3.19<br>(1.54)<sup>1</sup> | NA | 7.23<br>(4.79) | NA |
| PANSS total | NA | NA | 54.65<br>(10.63) | NA | 75.79<br>(10.44) | NA |
| PANSS negative | 16.69<br>(4.53)<sup>2</sup> | NA | 14.61<br>(4.19) | NA | 20.09<br>(5.24) | NA |
| PANSS positive | 20.75<br>(5.44)<sup>2</sup> | NA | 11.31<br>(3.13) | NA | 18.51<br>(4.37) | NA |
| Illness duration (months) | 8.87<br>(6.68) | NA | 54.38<br>(83.24) | NA | 63.13<br>(68.87) | NA |

**Note:** The table displays means and standard deviations of demographic (age, education and sex) and clinical information. Clinical symptoms were measured using the Scale for the Assessment of Negative Symptoms (SANS)<sup>37</sup>, the Scale for the Assessment of Positive Symptoms (SAPS)<sup>38</sup>, and the Positive and Negative Syndrome Scale (PANSS)<sup>39</sup>. Social functioning was measured using the Personal and Social Performance scale (PSP)<sup>40</sup>. NA = data not available; SCZ = patients with schizophrenia, HC = healthy controls.
<sup>1</sup> Scores calculated from the PANSS scores using the conversion formula provided in Van Erp et al. (2014).
<sup>2</sup> Scores calculated from the SANS/SAPS scores using the conversion formula provided in Van Erp et al. (2014).

### Voice recordings

The dataset included a total of 1843 recordings of individuals with schizophrenia (mean recording length = 17.5 seconds (sec), sd = 15.9sec) and 2009 recordings of control participants (recording length = 18.5sec, sd = 12.8sec). Voice recordings were collected using the Animated Triangles Task (Abell et al., 2000). The task is generally used to measure theory of mind (ToM) and involves twelve video clips representing an interaction between animated geometrical shapes (triangles). The participants were asked to provide an interpretation of what was going on in each animation and their answers were audio-recorded. A detailed description of the task and its validity for assessing speech production is included in the **SM2**. All recordings were pre-processed to remove background noise and interviewer speech when present. A full description of the process and of the extracted acoustic features is available in the **SM3**.

### Feature Sets

We used two different acoustic feature sets. The first feature set “Covarep” includes 434 acoustic features of rhythm, duration, voice quality and spectral and glottal properties of voice extracted using (see S3) the open-source library Covarep for Matlab (Degottex et al., 2014) and the open-source script syllablev2 for Praat (de Jong & Wempe, 2009; Quené et al., 2011), see SM3 for more details. The second feature set “eGeMAPS” includes 87 features extracted using the open-source feature extraction toolkit openSMILE 3.0 (Eyben et al., 2010). The choice of the feature sets was motivated by different reasons. First, all scripts and libraries used for feature extraction are openly available and thus allow for replicability of results. Second, these feature sets are largely used in previous literature (De Boer et al., 2023; Oomen et al., 2022a; Parola, Simonsen, et al., 2023; Rybner et al., 2021; Siriwardena et al., 2021), and thus allow comparability with previous studies. We scaled all features in the training set using min-max normalization to ensure uniform scaling without losing information or altering the variations in value ranges (Singh & Singh, 2020). The parameters determined for normalizing the training data were preserved and applied to the test dataset to prevent any potential information leakage between the two datasets.

### Data Splitting

Each dataset (DK, GE, CH) was partitioned into a training set and a test set, stratified by participant ID; all data points, i.e. each recording, for a given participant ended up in the same partition. The test set size was held constant (n = 32) across the language to allow comparability. We also ensured the test set included an equal number of participants from each diagnostic group (total number of voice recordings differed in some cases - see Table SM4_A), and were balanced for gender, to support more robust out-of-sample performance. A graphical representation of the data splitting procedure and the ML pipeline is provided in Fig. 1.

**Figure 1.**
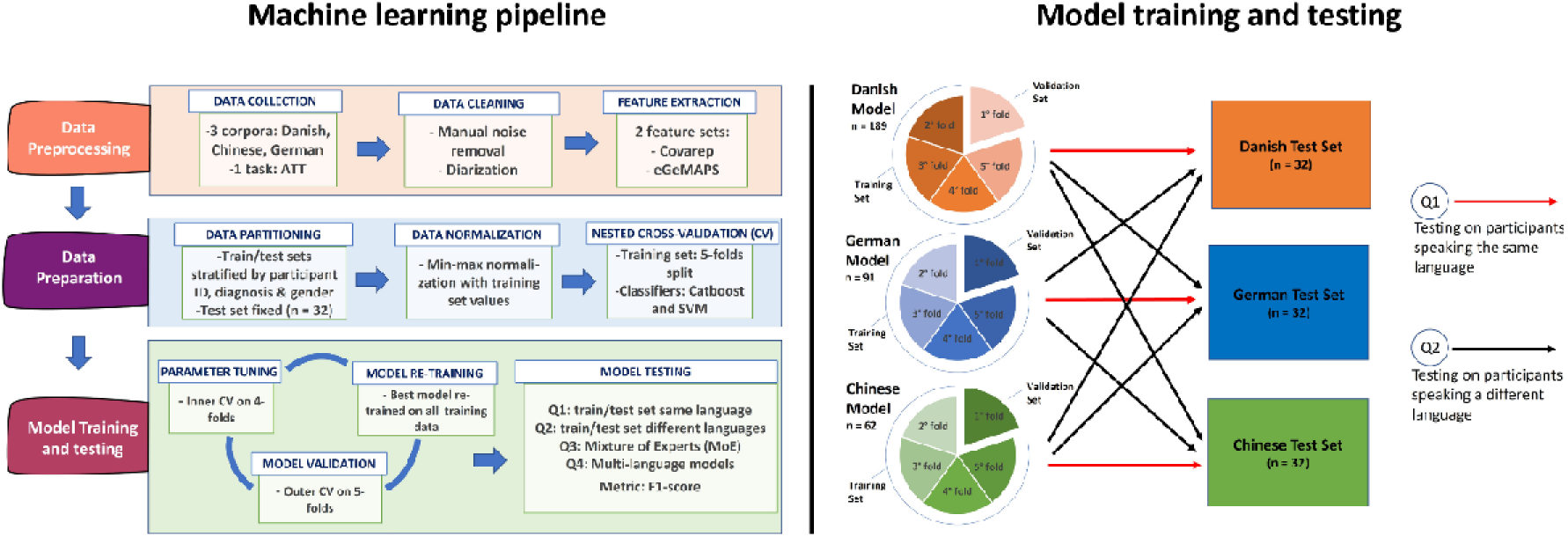
Overview of the machine learning pipeline (left plot) and the training and testing procedure (right plot).

### Machine learning models

We use two ML architectures. **Support Vector Machine** (**SVM**) is frequently employed in binary classification tasks and has been shown to often outperform other models, such as neural networks and Naive Bayes, in scenarios with limited training data and a large feature set. SVM performance is determined by three hyperparameters: γ (gamma, which determines the margin’s curvature between classes), and C (which dictates the allowable error margin).

**Gradient Boosted Decision Trees (GBDT**) is a very powerful machine learning model that combines multiple decision trees to improve overall model accuracy. Unlike traditional decision trees that are built independently, GBDT builds trees sequentially, where each new tree is trained to correct the errors made by the previous trees. GBDT is highly effective for a variety of tasks, offering flexibility, accuracy, and robustness to overfitting. We implemented GBDT using CatBoost, a recently introduced fast, scalable and high performance GBDT library (Prokhorenkova et al., 2018). Since our problem is binary classification, the GBDT was built by optimizing the log-loss. The main hyperparameters of GBDT are: a) the shrinkage parameter, b) the number of gradient boosting iterations (i.e. the number of decision trees), and c) the depth of the trees.

The choice of the classifiers was motivated by different reasons. First, SVM and decision trees are two widely employed classifiers and this allows comparability with previous studies (see table SM4_A). Second, we aimed to assess whether the computational complexity of the classifiers employed might affect the generalization performance (e.g. in terms of overfitting to the training set) of the models. SVM uses a regularization term (C) and is generally less prone to overfitting, especially when the number of dimensions (or features) is greater than the number of samples (as in this study); CatBoost instead can fit more complex relationships between features and target variables, making it more susceptible to overfitting, especially when the number of features is high relative to the number of observations (Berisha et al., 2021). By comparing SVM and CatBoost, we can assess whether the computational complexity of the employed classifier has an impact on overfitting and models’ generalization performance.

### Model training and cross-validation procedure

To answer Q1 and Q2, three models were trained (one for each language). We implemented a nested cross-validation (CV) approach, with an inner and an outer CV loop. We fine-tuned all mentioned hyper-parameters within the inner CV loop, whereas the outer CV loop is utilized to estimate the model’s generalization ability and to identify the best model (Cawley & Talbot, 2010; Sechidis et al., 2021). We chose a subject-wise CV approach, ensuring that the data for training, testing, and validation are from distinct individuals. This method - also known as speaker-independent CV - avoids potential data leakage across training, testing, and validation sets. Further, we set each group in our model to include individuals from both genders, maintaining a balanced gender ratio and avoiding models biased towards either gender. We employed a 5-fold group-wise CV, with each group comprising at least one male and one female subject (covering the various languages). Such balanced grouping is crucial; for instance, if the validation set in the inner CV loop only consists of female recordings, it could skew hyper-parameter tuning towards configurations that only benefit females, potentially underperforming for male subjects. The inner CV loop is dedicated to selecting hyperparameters that enhance a specific evaluation metric, which in our study is the F-measure. These chosen hyperparameters are then evaluated in the test set of the outer CV. Out of the five models from the outer CV (one per fold), we select the model with the highest F-measure for final use, which is then retrained with all the available data.

### Mixture of expert models

To answer Q3, we built three Mixture-of-Experts (MoE) ensemble models for the different languages. A MoE is an ensemble of models (experts) whose predictions are combined using a weighted sum to produce the overall prediction where the weights dynamically depend on the input. Initially, it was introduced to combine different neural networks, but given the above definition, it provides an effective way to create ensembles of any type of models. In our scenario, the probability of predicting the diagnostic label of a new participant, given her set of acoustic features, is provided by the weighted mixture of the predictions produced by the experts that were trained in different corpora. The experts in our corpus are represented by the three mono-language models (one for each language) described in the section “Model training”. The MoE models are created by combining the predictions by two experts (one for each language, for example Danish and Chinese) to predict the diagnosis of new participants speaking a third language (for example German).

### Multilanguage models

To answer Q4, multilanguage models were created by pooling datasets from different languages. For allowing comparability with the mono-lingual datasets, for each combination of languages (e.g. Danish and German) we created two pooled datasets, each with the size of the original monolingual dataset. For example, for the combination of Danish and German we created a) a Danish-German dataset with the size of the original Danish dataset, but with half data from the Danish dataset and half from the German dataset, and b) a German-Danish dataset with the size of the original German dataset but with half data from the German dataset and half from the Danish dataset. After the creation of the multilanguage training datasets, the multilanguage models were trained using the same procedure described in the “Model training” section.

### Testing procedure

Q1. The models were trained on each mono-lingual dataset (DK, GE, CH) using the procedure described in “Model training” section and then tested on new participants speaking the same language

Q2. The models were trained on each mono-lingual dataset (DK, GE, CH) and then tested on new participants speaking a different language

Q3. The predictions from two mono-lingual models (i.e. experts) were combined in a MoE ensemble model using the procedure described in the “Mixture of Expert models” section, and then tested on new participants speaking a different language.

Q4. The multi-language models were trained using the procedure described in the “Multilanguage models” section, and then tested on new participants speaking a different language. A graphical representation of the training and testing procedure is provided in Fig. 1.

F1 scores - a measure fit to scenarios where the groups to be classified have a different size - are computed as the harmonic mean of precision (the ratio of true positive instances among those identified as positive by the model) and recall (the ratio of true positive instances correctly recognized out of the overall actual positive instances). We calculated the F1-score for each class (patients and controls) and then computed the weighted average of F1 scores. To facilitate comparability with previous studies, we also report precision, recall and accuracy measures (see **SM5**).

## Results

**Q1:** The performance of the models is summarized in **Fig. 2** and **SM5**. Overall, the models trained and tested on the same language reported a high performance with an F1-Score between 0.60 and 0.84 comparable to state of the art results. Although a particular classifier or feature set may perform better on a particular training/test set (for example SVM and Covarep on the Danish corpora), there is no clear superiority of one classifier or feature set over another.

**Figure 2.**
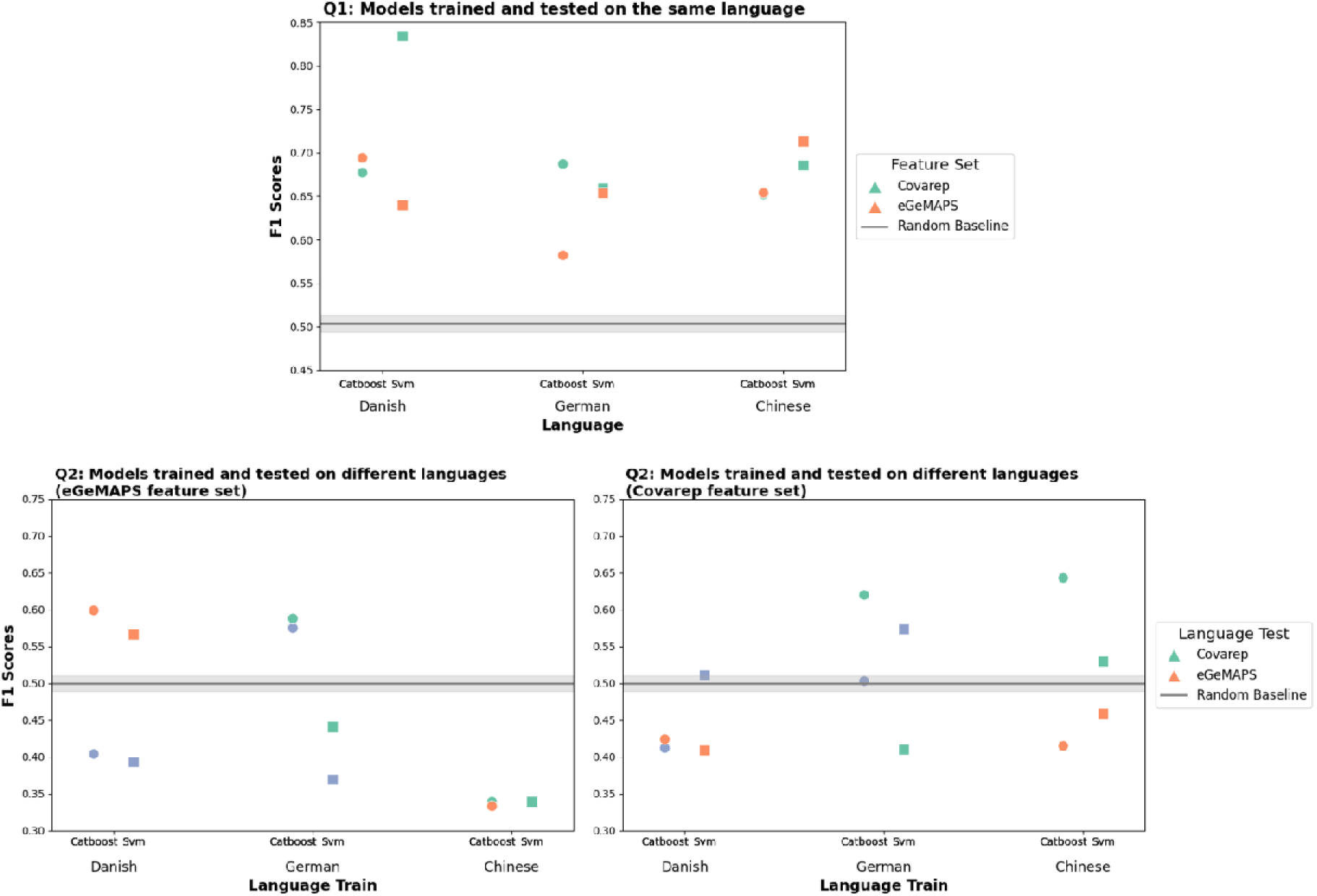
Test performance (F1 score) of models trained and tested on the same language (top plot, Q1) and trained and tested on different languages (bottom plots, Q2) for the eGeMAPS (left bottom plot) and Covarep (right bottom plot) features sets and the two classifiers (CatBoost and SVM) employed.

**Q2:** The performance of the models is summarized in **Fig. 2** and **SM5**. Overall, the models trained on a language (for example Danish) and tested on a different language (for example German) showed a low performance close to chance level with a F1-Score between 0.30 and 0.60.

**Q3:** The performance of the models is summarized in **Fig. 3** and **SM5**. Overall, MoE models showed a low performance close to or below chance level, with a F1-Score between 0.30 and 0.60.

**Figure 3.**
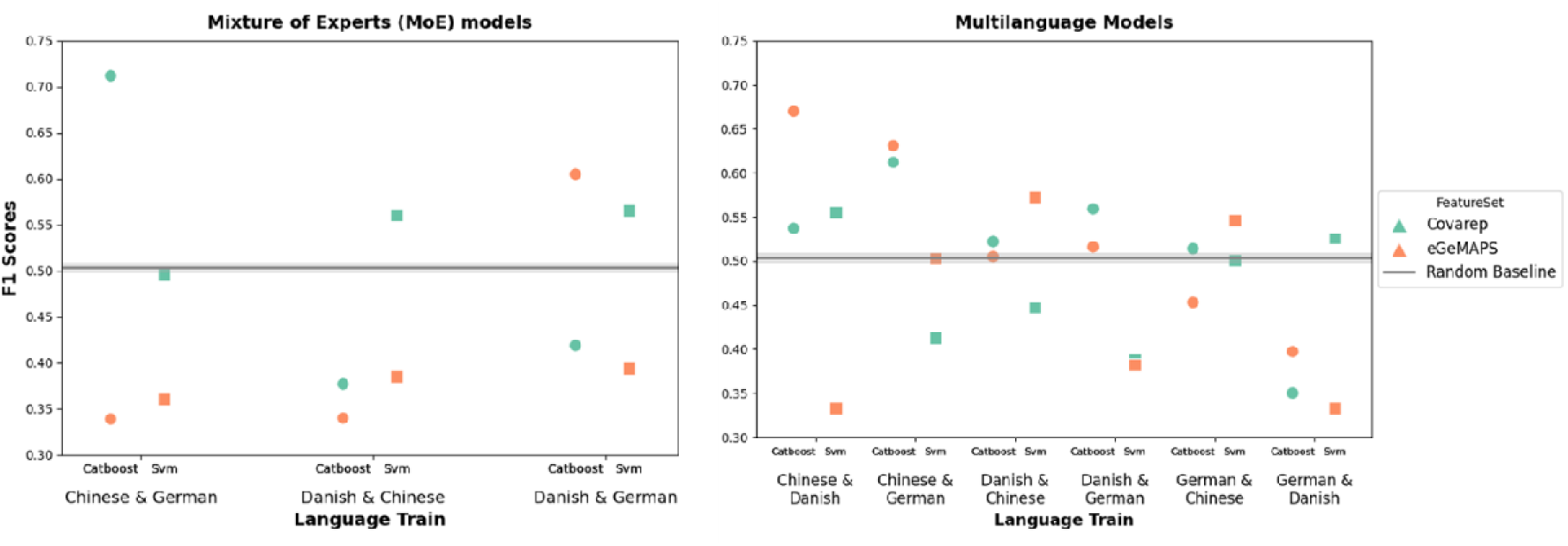
Test performance (F1 score) of mixture of experts (MoE, plot on the left) and multi-language models (plot on the right), for the two classifiers (eGeMAPS and Covarep) features set and the two classifiers (CatBoost and SVM) employed.

**Q4:** The performance of the models is summarized in **Fig. 3** and **SM5**. Overall, multi-language models showed a low performance close to chance level, with a F1-Score between 0.30 and 0.60.

### Overall results and model comparison

Overall, there is a clear drop of performance from models trained and tested on participants speaking the same language (F1 scores ∼ 0.70 - 0.80), compared to any other model (F1 scores, ∼ 0.50, see **Fig. 4**). MoE and multilanguage models might seem to show a slight increase in performance compared to models trained and tested on participants speaking a different language, but they do so in unsystematic ways, e.g. only for a specific classifier or feature set. Furthermore, when trained used the eGeMAPS feature set, some MoE models seem to be biased by one of the two experts that was overconfident in categorizing all the new participants either as patient or controls (see SM5) and resulting in a model performance lower than chance (F1 score = 0.33).

**Figure 4.**
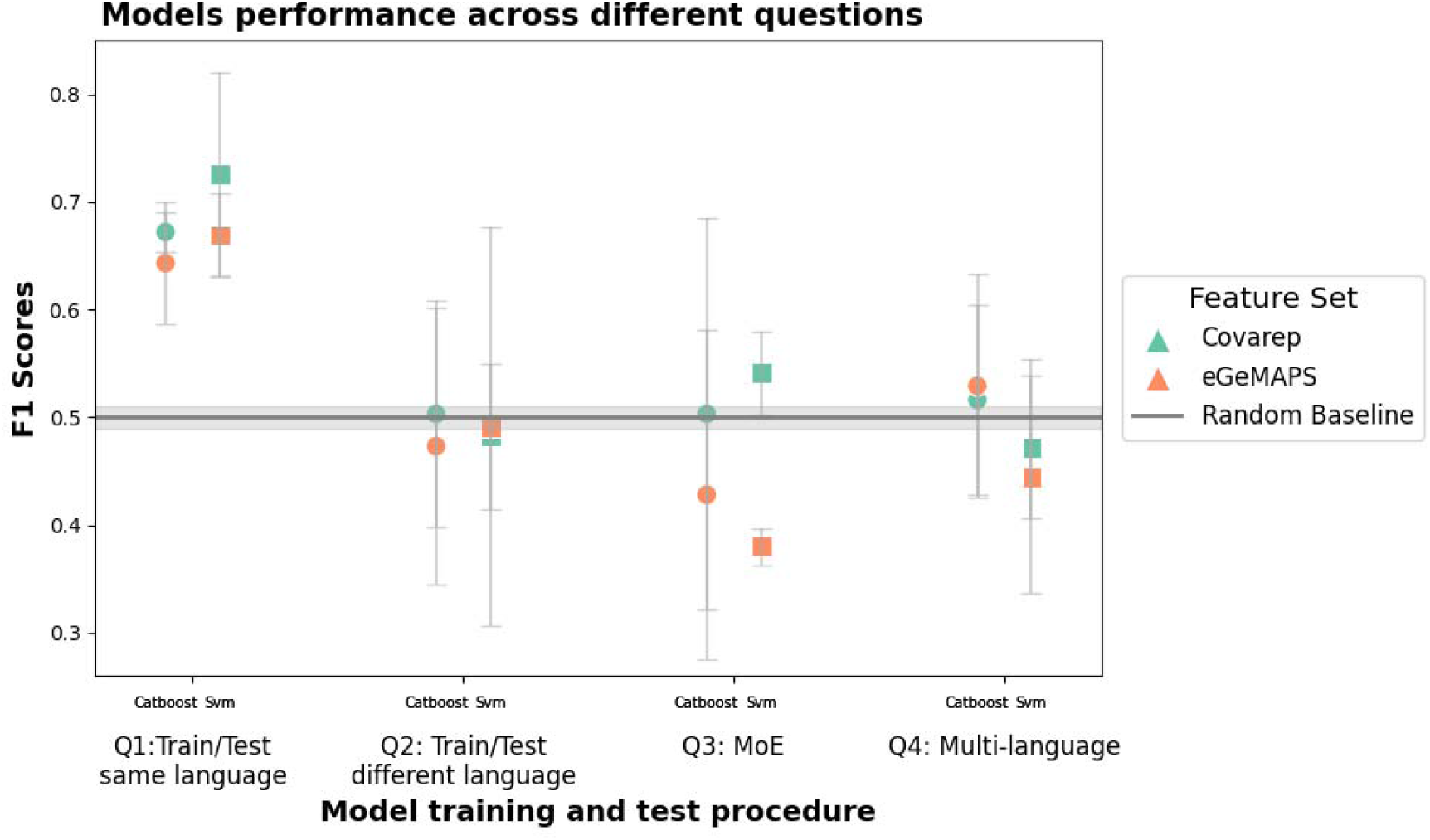
Mean test performance (F1 score) and standard deviation across the four different model categories (model trained and tested on the same language, model trained and tested on different languages, MoE models, multilanguage models) for the two feature sets (eGeMAPS and Covarep) and the two classifiers (CatBoost and SVM) employed.

### Performance in the training and test set

The performance of the models is summarized in **Fig. SM5_A** and **SM5_B**). For what concerns the Catboost classifier (see **Fig. SM5_A** and **SM5_B**), we see clear signs of overfitting in the training dataset with a very high classification performance (F1 scores ∼ 0.95), while performance in the test is much lower (F1 score ∼ 0.70).

For what concerns the SVM classifier, (**Fig. SM5_A** and **SM5_B**) the performance in the training and test sets is comparable, with a performance in the training sets only slightly higher than performance in the test sets.

## Discussion

### Aim

We systematically investigated the generalization performance of voice-based machine learning models predicting schizophrenia diagnosis, to assess their robustness and reliability in light of their clinical applicability. We tested how well these models generalize to different participants from the same study (Q1), to a different language (Q2), and whether generalizability could be improved by combining predictions from different monolingual models (Q3) or using multilanguage models (Q4).

### Main results

Overall, our results suggest that the models’ generalizability is limited. When models were tested on different participants from the same study (Q1), the performance was generally good and aligned with state-of-the art results (F1 scores ∼ 0.70 – 0.80). It is noteworthy that the performance in the training set was extremely good, especially for the high-performing classifier Catboost (F1 score ∼ 0.95), with clear signs of overfitting despite the rigorous cross-validation procedure we adopted. While the performance was tempered in the test set (F1 score ∼ 0.70 – 0.80), this indicates how high performing classifiers trained on small samples and with high-dimensional feature sets may reach excellent performance which however does not generalize well on future unseen patients. Further, the models’ performance on participants from a different study speaking a different language (Q2) was considerably lower, and at or below chance level on average (F1 scores ∼ 0.50). A good performance of a model trained on patients from a study was not a strong indicator of performance on future patients speaking a different language.

### MoE and multilanguage models

We then evaluated whether using training data with multiple languages, either in a Mixture of Expert (MoE) monolingual models (Q3) or in multilingually trained models (Q4), could improve generalizability to new languages. Our results showed that MoE or multilanguage models may show a better generalization performance compared to monolingual models (F1 score up to 0.60); however, it did so only for specific combinations of language, features and classifier, which questions how robust and generalizable the improvement might be. Indeed, on average, the performance of MoE and multilanguage models is not considerably better than that of monolingual models, and still close to chance level. Crucially, by exploring cross-linguistic generalizability we found that some of the acoustic features characterizing patients with schizophrenia in one study were shared by all participants (including controls) in another study (e.g. see the model in **Fig. SM5_C** classifying all test participants as patients). This suggests that transferring a voice-based ML model of schizophrenia across different studies and languages, even with multilingual training, is non-trivial and far from achieved in the field.

### Effects of sample size and features sets

A larger sample size seemed to matter only to improve generalizability to new participants from the same study (the larger dataset had better performance for Q1), but not to new languages (Q2-Q4). This does not exclude, however, that much larger sample sizes might be required to achieve better generalizability. Similarly, no feature set seemed clearly better than the other and the acoustic features used by the different models varied considerably.

### Why is generalization challenging?

We consider three possible reasons for this lack of generalizability: linguistic, individual and contextual heterogeneity. First, the differences between languages might have an impact on the way schizophrenia and its symptoms relate to acoustic features. Voice patterns are known to be affected by the phonological and morpho-syntactical structure of the language spoken (e.g. Çokal et al., 2019; Palaniyappan, 2021) and this variability could have contributed to reduce the models’ generalizability across languages. While the models did not generalize better to new languages of the same language family (e.g. Danish and German) than to a different language family (e.g., Danish and Chinese), there are still enough typological differences within the same family that could explain the lack of generalizability (Trecca et al., 2021). Second, participants’ heterogeneity might affect generalizability. Indeed, heterogeneity is widespread in schizophrenia (Dickinson et al., 2018; Gratton & Mittal, 2020) and patients with very different clinical and socio-demographic profiles, and corresponding vocal patterns are grouped under the same diagnostic category (see Cohen, Cox, Le, et al., 2020b; Oomen et al., 2022). In our corpus, we tried to limit differences between participants in different studies – e.g. in terms of symptom severity and inclusion criteria. Yet, other differences might have been present: in pharmacotherapy and other interventions, in time of onset, in cognitive profiles, in more specific symptom dimensions, etc. Third, we cannot exclude that subtle but important differences between studies in the recruitment process, as well as in the speech task used and administration procedure, might have also affected vocal patterns. Even keeping constant the speech task and recording conditions across studies (see **SM3**), we have observed a low consistency and high variability in which acoustic features were most relevant for the different models. This suggests that the models might not be capturing diagnosis- or symptoms-specific markers of schizophrenia, but rather learning heuristics to identify participants’ diagnosis based on subtle acoustic features which do not generalize well, even if the sociodemographic and clinical features of participants, and the recording conditions, are comparable across studies. Indeed, if the models are not capturing the information which distinguishes patients from controls and reflects the psycho-biological mechanisms of the disorder, predictions for new, unseen patients may lack accuracy.

This problem has already been highlighted in previous literature on speech markers. Voleti et al. (2020) found that reported accuracy in clinical speech marker studies diminishes as the sample size increases, indicating that studies with smaller samples tend to provide an excessively optimistic assessment of accuracy, mainly because of overfitting of models trained on large sets of features. Further, the problem of generalizability of clinical prediction models is not limited to speech markers but also affects models predicting outcomes of antipsychotic treatment (Chekroud et al., 2016, 2024) and extends to other candidate biological markers such as neuroimaging and genetic markers (Chekroud et al., 2021), posing a serious obstacle to clinical applicability.

To deal with these and other sources of heterogeneity, and understand the limits of generalizability of speech biomarkers, much larger datasets are needed, with much more fine-grained information on clinical and sociodemographic aspects and dedicated efforts to capture heterogeneity in languages, participants, and contexts.

### How to improve generalization?

First, our data suggests that the first necessary step is to thoroughly document and test the generalizability of voice-based ML models. As we have shown, and in line with previous studies, within-study out-of-sample validation is not a reliable indicator of generalizable findings (Rybner et al., 2021; Voleti et al., 2020). The relatively small size and homogeneity of the datasets involved in previous studies may have not provided a realistic estimate of voice-based ML models performance, where the findings would likely apply and where they would not (Parola, Lin, et al., 2023; Parola, Simonsen, et al., 2020b, 2023). Any realistic assessment of generalization performance thus requires larger multi-language datasets with data from multiple studies (see Rocca & Yarkoni, 2021). These datasets should strive to represent as much variability in language spoken, clinical, cognitive and socio-demographic profiles, as well as in speech tasks and recording settings.

Such datasets would further allow us to identify which acoustic features generalize better and in which contexts thus providing insights on the underlying psycho-biological mechanisms. Not least, more recent development in ML models - such as time sensitive algorithms, large audio models and transformer architectures (see Babu et al., 2021; Bain et al., 2023), and multimodal models (e.g., Wolf et al., 2023) - require such large datasets to be trained or even just fine-tuned. Indeed, when used on clinical datasets for purposes similar to those in this paper, their efficacy was found limited (Hansen et al., 2023). In this respect, using synthetic data or augmented clinical datasets with data from healthy individuals can partially remedy the data shortage and small sample sizes in the field (e.g. Mehta et al., 2024).

Finally, this work focused on predicting diagnostic groups, while predicting more fine-grained clinically relevant features can enhance generalization and clinical applicability of voice-based ML models. In this respect, one of the crucial challenges is to identify which clinically relevant features are robust across context, and accurately assess the performance of models trained with those features (e.g. Bilgrami et al., 2022; Tan et al., 2021). Another crucial challenge is to predict how voice-related symptoms evolve and interact with each other over time, and recent approaches combining remote collection of speech markers and clinical information via ecological momentary assessment (EMA) and digital devices offer a promising venue for broadening our understanding of mental disorders (Abbas et al., 2022; Cohen, Cox, Masucci, et al., 2020; Fried et al., 2023; Miller et al., 2022). If the ultimate goal is clinical applicability, the ability to predict longitudinally and ecologically fine-grained clinical features from interpretable acoustic models could more realistically assist practitioners in monitoring symptoms over time, in identifying the most effective treatments or provide them with prognostic indicators on, for instance, the probability of relapse.

## Data Availability

The anonymized acoustic features extracted described in the paper are available here: https://osf.io/h4wj6/

## Acknowledgments

A.P. was supported by a Marie Skłodowska-Curie Actions – H2020-MSCA-IF-2018 grant (ID: 832518, Project: MOVES). K.K. has been supported by Japan Society for the Promotion of Science (JSPS) (PE 07550). The study was supported by seed funding from the Interacting Minds Center (Aarhus University).

