## Supplementary Material for "Vocal markers of schizophrenia: assessing the generalizability of machine learning models and their clinical applicability"

The supplementary materials contain the following sections:

- SM1: Description of the voice recordings corpus

- SM2: Animated Triangles Task (ATT)

- SM3: Audio preprocessing and feature extraction

- SM4: Review of previous ML studies and ML pipeline information

- SM5: Results

- SM6: Feature importance

- SM7: Comparison of models trained and tested on similar or different language families

**S1 – Description of the voice recordings corpus**

**Danish corpus**

The patient and control samples for the present study were collected within three consecutive studies recruiting patients at the same clinical location, i.e., at OPUS - Clinic for people with schizophrenia (SCZ), Aarhus University Hospital Risskov, in the period from 2009 to 2018^1–3^.

The patients had been diagnosed by experienced psychiatrists according to ICD-10 criteria. Exclusion criteria were a history of neurological disorder, severe head trauma, or substance abuse problem according to ICD-10. Patients were excluded if they did not understand spoken Danish sufficiently to understand the testing procedures or if they had an estimated premorbid IQ below 70 based on their history.

All patients with schizophrenia, except the sample reported in Veddum et al. (2019)^3^, were interviewed with the Scale for Assessment of Negative Symptoms (SANS), the Scale for Assessment of Positive Symptoms (SAPS), and the Personal Social Performance Scale (PSP), a measure of social functioning in schizophrenia.

**Control groups**

Patients and non-clinical controls were matched on age, sex, and level of education (except sample 3, matched only on age and sex). Control participants did not have a history of neurological or mental illness (and neither had their first-degree relatives), severe head injury or drug- or alcohol dependence according to ICD-10 criteria. Demographics and social functioning of the patients and their matched controls are summarized in **Table 1.**

**IQ**

Verbal intelligence was estimated from two subtests (Vocabulary and Similarities) from WAIS-III (Wechsler Adult Intelligence Scale, Third edition^4^). The two subtests were chosen based on their high correlation with the verbal WAIS-III IQ-score.

**Ethics**

The participants received written and oral information about the project, and written informed consent was obtained before inclusion. The study was approved by The Central Denmark Region Committees on Biomedical Research Ethics (Ref: M-2009–0035; Ref: 2007-58-0010) and the Danish Data Protection Agency. The project complied with the Helsinki Declaration of 1975, as revised in 2008.

**Chinese corpus**

The patient and control samples were collected within two consecutive studies recruiting patients at the same clinical location, i.e., Renmin Hospital of Wuhan University^1,5^, and a part of the data (n = 41 participants) has not yet been published.

Patients met the diagnostic criteria for schizophrenia according to ICD-10 and were diagnosed by psychiatrists.

Exclusion criteria were a history of severe head trauma or neurological illness or if they had a substance abuse problem according to the ICD-10. Patients with an estimated premorbid IQ below 70 based on previous history or who were unable to understand spoken Chinese well enough to understand testing instructions were also excluded. All of the Chinese patients were of Han Chinese ethnicity.

Patients were interviewed with the Scale for Assessment of Negative Symptoms (SANS), the Scale for Assessment of Positive Symptoms (SAPS), the Personal Social Performance Scale (PSP).

**Control group**

There were no differences between patients and non-clinical controls on age and sex at group level. The exclusion criteria for the healthy subjects were the same as for patients. In addition, healthy control subjects were excluded if they, or a first-degree relative, met any psychiatric diagnosis according to ICD-10.

**IQ**

Verbal intelligence was estimated from Vocabulary subtests from WAIS-III (Wechsler Adult Intelligence Scale, Third edition)^4^. The subtest was chosen based on its high correlation with the verbal WAIS-III IQ-score^4^.

**Ethics**

All participants received written and spoken information about the project, and written consent was obtained. The project was approved by the Ethics committee of Renmin Hospital of Wuhan University and the Institutional Review Board of the Institute of Psychology, the Chinese Academy of Sciences. The authors assert that all procedures contributing to this work comply with the ethical standards of the relevant national and institutional committees on human experimentation and with the Helsinki Declaration of 1975, as revised in 2008.

**German corpus**

The patient and control samples for the present study were collected within three consecutive studies recruiting patients at the same clinical location, i.e., the University of Muenster and the LWL-Hospital Muenster^7–9^ (from 2005 to 2017).

The patients had been diagnosed with schizophrenia by experienced psychiatrists using the Structured Clinical Interview for DSM-IV SCID-I. Psychopathology was assessed with the Positive and Negative Syndrome Scale (PANSS).

Patients with any history of other psychiatric disorders, neurological disorders, serious head injury, alcohol or illegal drug abuse, or insufficient knowledge of the German language were excluded from the study.

**Control group**

Healthy controls (HC) were matched with patients for age, sex and education. Healthy participants with no history of Axis-I DSM-IV diagnoses (SCID), illegal drug use, alcohol abuse or addiction, or neurological disorders served as a control group. Subjects with any first-degree relatives with a history of mental disorders were excluded from the study.

**Ethics**: After hearing a complete description of the study, written informed consent was obtained from all participants. The studies were approved by the Ethics Committee of the State Chamber of Physicians Westphalia-Lippe and the University of Muenster and have been carried out in accordance with the Declaration of Helsinki.

**S2 – Animated Triangles Task**

Voice recordings were collected using the Animated Triangles Task^11,12^. The task is generally used to measure theory of mind (ToM) and involves between eight to twelve video clips representing an interaction between animated geometrical shapes (triangles). In some of the clips the two triangles are moving randomly and unintentionally (e.g., bouncing off walls) (4 clips), and in the other clips the triangles are interacting intentionally to influence the mental state of one another (e.g., a larger triangle trying to convince a small one to leave a closure) (4 clips) or merely performing an activity alone or together (4 clips). The duration of each animation is approximately 40 seconds. The participants were asked to provide an interpretation of what was going on in each animation and their answers were audio-recorded. Not all studies used the 4 pure action clips.

As shown in a recent meta-analysis^13^, free speech tasks and dialogic tasks with social requirements, compared to constrained speech (e.g., reading), had significantly larger effects both in contrasting patients and controls and in assessing symptomatology. The Animated Triangle Task is thus a more suitable task (compared to, e.g. a constrained speech task) for assessing whether voice patterns can be a marker of schizophrenia and comparing them across languages^14^.

**S3 – Voice recordings preprocessing and features extraction**

All audio recordings were carefully listened to in order to identify potential audio quality issues that would lead to the exclusion of the recording. All audio recordings were then manually timecoded, to identify the segments corresponding to the participants’ video description, thus excluding instructions, the prompts (e.g., “Can you say anything more about that?”) as well as questions, backchannels such as (‘hmmm’, or ‘ok’, etc.) provided by the examiner. Once the relevant audio segments had been identified, we proceeded to clean the audio. Background room noise, reverberation and hum were removed from the audio recordings using iZotope RX 6 ElementsTM. Long-term average spectra for each recording were inspected for possible noise artefacts and further cleaned if any were found.

We first extracted features every 10 milliseconds using Covarep for Matlab^17^. The feature set comprises a wide range of acoustic features, including voice quality, spectral and glottal features of voice. Mean, median, standard deviation, interquartile range (IQR) and median absolute deviation (MAD) were calculated for each of these measures using R^18^. This yielded a total of 361 features.

We then extracted measures of rhythm and duration for the full relevant segment (e.g., one video description) using the open-source script syllablev2 for Praat^15,16^. Indeed, in line with a previous study and previous meta-analyses, duration and rhythm measures were identified as characterizing speech of individuals with schizophrenia, and were thus included in our features set. Further, to complement the voice quality features extracted with Covarep, we extracted additional voice quality measures including jitter and shimmer features (for a more detailed description see **Table S3_A**) which revealed to be important for detecting other speech disorders (e.g. Cummins et al., 2015). This yielded 73 measures of pitch, duration and voice quality, for a total of 361 features. The entire set thus included 434 acoustic features.

Finally, we extracted the standardized voice feature set eGeMAPS with the open-source software OpenSmile (Eyben, 2015; Eyben et al., 2010, 2016). The feature set contains 87 features (see Table S3_B).

**Table S3_A. Covarep feture set**

| **Spectral and glottal Acoustic Features** | **Description** |
| --- | --- |
| Formants frequency (F1- F2 -F3 -F4 -F5) | Range of frequencies in which there is absolute or relative maximum in the sound spectrum. The frequency at the maximum is the formant frequency. |
| Relative amplitude (H1H2) | Amplitude of the first harmonic relative to the second harmonic. Associated with breathiness. |
| Harmonics to Noise Ratio (HNR) | Ratio of harmonics to inharmonic (spectral components which are not a whole number multiple of F0) components. Associated with hoarseness. |
| Normalized Amplitude Quotient (NAQ) | Parametrization of the glottal closing phase using two amplitude-domain measurements from waveforms estimated by inverse filtering. Associated with breathiness. |
| Quasi-Open-Quotient (QOQ) | Relative open phase duration of a glottal cycle. Associated with breathiness. |
| Maxima Dispersion Quotient (MDQ) | Sharpness of the glottal excitation. Associated with tense-lax properties of the voice and relatedly with breathiness. |
| Parabolic Spectral Parameter (PSP) | A quantification of the glottal volume velocity waveform. Associated with breathiness and tense phonation. |
| Harmonic Richness Factor (HRF) | Ratio between the sum of the amplitudes of harmonics, and the amplitude at the fundamental frequency, quantifying the amount of harmonics in the magnitude spectrum of the glottal source. |
| MCEP | Transforming the spectrogram into a Mel spectrum using the Mel scale filter bank, followed by cepstrum analysis. |
| HMPDM- HMPDD | Harmonic Model Phase Distortion Mean and Harmonic Model Phase Distortion Deviation offer a flexible representation of the glottal source based on short-term phase distortion statistics. |
| Peak-to-RMS | The peak-to-RMS measure reflects a local loudness metric related to the waveform shape over a few pitch periods. |
| MFCC-deltas | These reflect the dynamic information of the spectrum envelope on a frame of the voice signal. |
| Srh | Summation of Residual Harmonic is a measure used to evaluate the harmonic structure of a voiced segment. This measure helps in distinguishing between different types of phonation (e.g., modal, breathy, and tense voices) by analyzing the residual harmonic content that remains in the signal after the primary harmonic components are accounted for. |
| Creak | These features reflect functions used to detect creaky voice (otherwise known as vocal/glottal fry) using novel acoustic features developed by Kane & Drugman (KD features) as well as previous acoustic features developed by Carlos Ishi and colleagues. Detection is done using artificial neural networks (see Degottex et al., 2014). |
| Rd | This parameter represents the glottal flow's shape and is used to describe the glottal waveform's relative deviation from an idealized waveform. It is a dimensionless quantity often used in glottal inverse filtering to capture the voice quality and characteristics of the speech signal. The Rd shape parameter of the Liljencrants-Fant (LF) glottal model is determined using the Mean Squared Phase (MSP) method based on MSPD2. |
| Peak Slope | The slope coefficient of a regression line fits to local peaks using wavelet analysis. |

| **Duration and voice quality features** |  | **Description** |
| --- | --- | --- |
| Fundamental frequency – F0 | Pitch median | Pitch mean reflects the mean frequency of vibrations of the vocal cords during vocal production across the linguistic unit analyzed (phoneme, word, sentence, or the entire speech sample). |
|  | Pitch variability (IQR) | Pitch variability indicates the mean magnitude of changes in pitch across the linguistic unit analyzed (phoneme, word, sentence, or the entire speech sample). |
| Speech production | Speech rate | Speech rate is defined as the number of words per time unit (second). |
|  | Percent time talking | Percent time talking represents the percentage of time the speech sample contained a pitch different from zero, i.e. speaking time, relative to the total time of the speech sample. |
|  | Duration of pauses | Mean pause duration indicates the mean duration of pauses across the entire speech sample. |
|  | Number of pauses | Mean number of pauses across the entire speech sample per time unit (second). |
|  | Duration of utterance | Median utterance duration indicates the median duration from beginning to end of the defined speech sample per time unit (second). |
|  | Number of utterances | Mean number of sentences across the entire speech sample per time unit (second). |
| Jitter |  | Jitter quantifies the frequency instability from one vocal cycle to the next, providing an indication of the regularity of vocal fold vibrations. |
| Shimmer |  | Shimmer measures the amplitude instability from one vocal cycle to the next, providing insights into the consistency of vocal fold vibration intensity. |

***Table S3_B***

**eGeMAPS Feature Set Summary**

| **Feature** | **Description** |
| --- | --- |
| Loudness | Perceptual loudness |
| Alpha Ratio | Ratio of spectral energy in 50-1000 Hz to 1-5 kHz |
| Hammarberg Index | Ratio of spectral energy in 0-2 kHz to 2-5 kHz |
| Slope of Regression Line (0-500 Hz) | Slope of the linear regression of the logarithmic power spectrum in the range 0-500 Hz |
| Slope of Regression Line (500-1500 Hz) | Slope of the linear regression of the logarithmic power spectrum in the range 500-1500 Hz |
| MFCC1 | Mel-frequency cepstral coefficient 1 |
| MFCC2 | Mel-frequency cepstral coefficient 2 |
| MFCC3 | Mel-frequency cepstral coefficient 3 |
| MFCC4 | Mel-frequency cepstral coefficient 4 |
| F0 | Fundamental frequency |
| Jitter (Local) | Frequency variation from cycle to cycle (jitter) |
| Shimmer (Local) | Amplitude variation from cycle to cycle (shimmer) |
| HNR | Harmonics-to-noise ratio |
| F0 Envelope | Envelope of the fundamental frequency contour |
| Voicing Probability | Probability of voicing |
| Voiced Segment Length | Length of voiced segments |
| Unvoiced Segment Length | Length of unvoiced segments |
| Voiced/Unvoiced Segment Ratio | Ratio of lengths of voiced to unvoiced segments |
| F0 Contour | Contour of the fundamental frequency |

**SM4 - Review of previous ML studies and ML pipeline information**

In this section we provide a table summarizing previous studies which used ML techniques and acoustic feature sets to classify patients with SCZ and controls or predict clinical symptoms of schizophrenia (**Table SM4_A**).

We also provide an additional table showing the exact number and demographic characteristics of participants for both groups (patients and controls) for the training and test sets for each language (**Table SM4_B**).

Table **SM4_A**. Previous studies using ML techniques and acoustic feature sets to classify patients with SCZ and controls or predict clinical symptoms.

| **Study** | **Aim** | **Sample** | **Classifier** | **Cross-validation** | **Acoustic features** | **Index and results** |
| --- | --- | --- | --- | --- | --- | --- |
| Rapcan et al. (2009)^[1]^ | Predict diagnosis | A sample of 57 participants, 39 patients with schizophrenia, and 18 control subjects | LDA | Cross-fold validation | Number of Pauses, Proportion of Silence, Total Recording Time, Total Length of Pauses (s). | Sensitivity (%) 72.64 75.21 72.36  Specificity (%) 78.63 83.62 85.47  Positive predictivity (%) 77.86 82.02 83.39  Negative predictivity (%) 74.24 77.40 75.75  Overall accuracy (%) 75.64 79.42 78.92  Area under the ROC curve 0.79 0.82 0.80 |
| Martinez-Sanchez et al. (2015) | Predict diagnosis | A sample of 80 participants,  45 patients diagnosed with schizophrenia  and 35 asymptomatic controls | LDA | NR | Task duration (s), Intensity (dB), Pause rate, Pitch Mean F0 (Hz), Pitch variability SD (Hz), F0 Range (ST), Syllabic dynamics, Prosodic peaks, Prosodic valleys, Intra-syllabic Trajectory (ST/s), Inter- syllabic Trajectory (ST/s), Phonation Trajectory (ST/s | Overall accuracy: 87.5% |
| Kliper et al. (2015) | Predict diagnosis | 42 participants, patients with schizophrenia (n=22) and healthy participants (n=20) | SVM | Leave-one-out | Spoken Ratio, Utterance Duration, Gap Duration, Pitch Range, Pitch variability, Power variability, Mean Waveform Correlation (MWC), Jitter, Shimmer | Overall accuracy: 76.19% |
| Puschel et al. (1998) | Predict diagnosis and symptoms | 90 participants, 45 acute hospitalized patients with schizophrenia, and 45 matched controls | LDA | NA | Total recording time, total length of utterances, number of pauses, mean energy per second, variation of energy per second and JO-contour. | Overall accuracy: 85.6%  3.6 % false positive  22.6 % false negative  After 2 weeks:  overall accuracy: 83.3%.  4.4 % false positive  28.9 % false negative  Symptoms:  Overall accuracy: 78.6%  After 2 weeks: 71.4%. |
| Cohen et al. (2020) | Predict symptoms | Participants (n = 121 total; 57 in Study 1 and 64 in Study 2) were stable  outpatients meeting definitions of SMI | Regularized regression | 10 analytic-folds | Computerized assessment of Affect from Natural Speech (CANS, 68 distinct features) + GeMAPS | 0.85% - 0 .92% |
| Thair et al. (2019) | Predict diagnosis and symptoms | 77 participants, 51 outpatients diagnosed with schizophrenia  and 26 healthy individuals | SVM, Random Forest, multilayer perceptron (MLP), Ensemble (bagging) | Leave-one-patient-out cross-validation | Matlab based features. | 70 – 81.5% |
| Espinola et al., (2021) | Predict diagnosis | 31 individuals over 18 years, 20 with diagnosis of schizophrenia, and 11  healthy controls. | MLP, logistic regression, random forest (RF), decision trees, Bayes net, Naïve Bayes, and SVM | 10-fold cross-validation | 33 Matlab-based parameters | 77% - 91% |
| de Boer et al., (2022) | Predict diagnosis and symptoms | 284 participants, 142 patients with a schizophrenia-spectrum disorder and  142 matched controls | Random forest | Leave-ten-out cross-validation | eGeMAPS | 0.86% |
| Agurto et al. (2020) | Predict CHR+ | 32 subjects at CHR of  Psychosis: five developed  psychosis (CHR+) within a period of 3 to 16 months. | SVM, Logistic regression | Nested 10-fold cross validation | Praat-based parameters | 0.84% |

**Table SM4_B.** Demographic characteristics of patients with schizophrenia and healthy controls (HC) for the training and test sets for each language

| **Training Sets** | | | | | | |
| --- | --- | --- | --- | --- | --- | --- |
| **Corpus** | **Danish** | | **German** | | **Chinese** | |
| **Diagnosis** | SCZ  N = 89 | HC  N = 100 | SCZ  N = 45 | HC  N =46 | SCZ  N = 35 | HC  N = 27 |
| **N. of recording** | N =764 | N = 859 | N =447 | N = 450 | N = 214 | N = 275 |
| **Age** | 26.5  (8.82) | 26.4  (8.96) | 31.7  (9.92) | 33.2  (8.79) | 27.2 (7.25) | 29.7  (8.72) |
| **Education** | 26.1 | 26.8 | 32.18 | 33.4 | 25.71 | 28.38 |
| **Sex (n. of males and %)** | 52 (58%) | 58 (58%) | 31 (68%) | 30 (65%) | 20 (57%) | 16 (59%) |
| **Test Sets** | | | | | | |
| **Corpus** | **Danish** | | **German** | | **Chinese** | |
| **Diagnosis** | SCZ  N = 16 | HC  N =16 | SCZ  N = 16 | HC  N = 16 | SCZ  N = 16 | HC  N = 16 |
| **N. of recording** | N = 130 | N = 136 | N = 162 | N = 162 | N = 126 | N = 126 |
| **Age** | 29.03 | 24.07 | 30.38 | 32.73 | 30.53 | 31.9 |
| **Education** | 13.59 | 13.8 | 11.86 | 12.5 | 12.89 | 14.2 |
| **Sex (n. of males and %)** | 8 (50%) | 8 (50%) | 8 (50%) | 8 (50%) | 8 (50%) | 8 (50%) |

**SM5 – Results**

In this section we provide additional tables summarizing the test performance (F1 score) for the models trained and tested on the same language (Q1, , Table SM5_A) and the models trained and tested on different languages (Q2, Table SM5_A), the test performance (F1 scores) for the Mixture of Experts models (Table SM5_B) and multilanguage models (Table SM5_C), and all the relevant metrics (F1 score, Accuracy, Precision and Recall) for all the models trained and tested on the same language and different languages (Table SM5_D and Table SM5_E). We also provide additional figures showing the test performance (F1 score) in the training and test set for each of the 5 models trained in the 5-folds cross-validation procedure (Figures SM5_A and SM5_B). Finally, we provide additional figures showing the performance of MoE models and monolingual expert models and their confidence in classifying either patients or controls (Figures SM5_C to SM5_P).

**Table SM5_A.** *Test performance (F1 score) for the models trained and tested on the same language (cells on the diagonal) and the models trained and tested on different languages (out of diagonal cells), for the two features set (eGeMAPS and Covarep) and the two classifiers (SVM on the left side of the slash, Catboost on the right side) used.*

| **Covarep feature set** | | | |
| --- | --- | --- | --- |
| **Tested on →**  **Trained on ↓** | **Danish** | **German** | **Chinese** |
| **Danish** | Q1: 0.68/0.83 | Q2: 0.42/0.41 | Q2 0.41/0.51 |
| **German** | Q2: 0.62/0.41 | Q1: 0.69/0.66 | Q2: 0.50/0.57 |
| **Chinese** | Q2: 0.64/0.53 | Q2: 0.42/0.46 | Q1: 0.65/0.69 |
| **eGeMAPS feature set** | | | |
| **Tested on →**  **Trained on ↓** | **Danish** | **German** | **Chinese** |
| **Danish** | Q1: 0.70/0.64 | Q2 0.60/0.57 | Q2: 0.40/0.39 |
| **German** | Q2: 0.59/0.44 | Q1: 0.59/0.65 | Q2: 0.58/0.37 |
| **Chinese** | Q2: 0.34/0.34 | Q2: 0.33/0.83 | Q1: 0.65/0.71 |

**Table SM5_B.** *Test performance (F1 score) for the Mixture of Experts (MoE) models, for the two features set (eGeMAPS and Covarep) and the two classifiers (SVM on the left side of the slash, Catboost on the right side) used.*

| **Covarep feature set** | | | |
| --- | --- | --- | --- |
| **Tested on →**  **Trained on ↓** | **Danish** | **German** | **Chinese** |
| **Danish and German** |  |  | Q3: 0.42/0.57 |
| **Danish and Chinese** |  | Q3: 0.38/0.56 |  |
| **Chinese and German** | Q3: 0.71/0.50 |  |  |
| **eGeMAPS feature set** | | | |
| **Tested on →**  **Trained on ↓** | **Danish** | **German** | **Chinese** |
| **Danish and German** |  |  | Q3: 0.61/0.39 |
| **Danish and Chinese** |  | Q3: 0.34/0.39 |  |
| **Chinese and German** | Q3: 0.34/0.36 |  |  |

**Table SM5_C.** *Test performance (F1 score) for the multilanguage models, for the two features set (eGeMAPS and Covarep) and the two classifiers (SVM on the left side of the slash, Catboost on the right side) used.*

| **Covarep feature set** | | | |
| --- | --- | --- | --- |
| **Tested on →**  **Trained on ↓** | **Danish** | **German** | **Chinese** |
| **Danish and German** |  |  | **Q4: 0.56/0.39** |
| **German and Danish** |  |  | **Q4: 0.35/0.53** |
| **Danish and Chinese** |  | **Q4: 0.52/0.45** |  |
| **Chinese and Danish** |  | **Q4: 0.54/0.56** |  |
| **Chinese and German** | **Q4: 0.61/0.41** |  |  |
| **German and Chinese** | **Q4: 0.51/0.50** |  |  |

| **eGeMAPS feature set** | | | |
| --- | --- | --- | --- |
| **Tested on →**  **Trained on ↓** | **Danish** | **German** | **Chinese** |
| **Danish and German** |  |  | **Q4: 0.52/0.38** |
| **German and Danish** |  |  | **Q4: 0.40/0.33** |
| **Danish and Chinese** |  | **Q4: 0.51/0.57** |  |
| **Chinese and Danish** |  | **Q4: 0.67/0.33** |  |
| **Chinese and German** | **Q4: 0.63/0.50** |  |  |
| **German and Chinese** | **Q4: 0.45/0.55** |  |  |

**Table SM5_D.** *Metrics performance (F1 score, Accuracy, Precision and Recall) for the models trained and tested on the same language.*

| **Language** | **Feature**  **Set** | **Classifier** | **F1** | **Accuracy** | **Precision**  **SCZ** | **Precision**  **CT** | **Recall**  **SCZ** | **Recall**  **CT** |
| --- | --- | --- | --- | --- | --- | --- | --- | --- |
| Chinese | Covarep | Catboost | 0.65 | 0.66 | 0.62 | 0.72 | 0.8 | 0.52 |
| Chinese | Covarep | SVM | 0.69 | 0.69 | 0.67 | 0.71 | 0.74 | 0.64 |
| Chinese | eGeMAPS | Catboost | 0.65 | 0.66 | 0.62 | 0.74 | 0.82 | 0.5 |
| Chinese | eGeMAPS | SVM | 0.71 | 0.72 | 0.67 | 0.8 | 0.85 | 0.59 |
| Danish | Covarep | Catboost | 0.68 | 0.68 | 0.75 | 0.64 | 0.56 | 0.81 |
| Danish | Covarep | SVM | 0.83 | 0.84 | 0.92 | 0.78 | 0.74 | 0.93 |
| Danish | eGeMAPS | Catboost | 0.69 | 0.7 | 0.77 | 0.65 | 0.58 | 0.82 |
| Danish | eGeMAPS | SVM | 0.64 | 0.64 | 0.71 | 0.61 | 0.53 | 0.77 |
| German | Covarep | Catboost | 0.69 | 0.69 | 0.71 | 0.67 | 0.64 | 0.74 |
| German | Covarep | SVM | 0.66 | 0.66 | 0.68 | 0.64 | 0.6 | 0.72 |
| German | eGeMAPS | Catboost | 0.58 | 0.58 | 0.57 | 0.59 | 0.64 | 0.53 |
| German | eGeMAPS | SVM | 0.65 | 0.65 | 0.66 | 0.65 | 0.64 | 0.67 |

**Table SM5_E.** *Metrics performance (F1 score, Accuracy, Precision and Recall) for the models trained and tested on different languages.*

| **Lang.**  **Train** | **Lang.**  **Test** | **Feature**  **Set** | **Classifier** | **F1** | **Accuracy** | **Precision**  **SCZ** | **Precision**  **CT** | **Recall**  **SCZ** | **Recall**  **CT** |
| --- | --- | --- | --- | --- | --- | --- | --- | --- | --- |
| Chinese | Danish | Covarep | Catboost | 0.64 | 0.64 | 0.66 | 0.63 | 0.62 | 0.67 |
| Chinese | Danish | Covarep | SVM | 0.53 | 0.54 | 0.57 | 0.52 | 0.4 | 0.68 |
| Chinese | Danish | eGeMAPS | Catboost | 0.34 | 0.51 | 0.51 | 0.0 | 1.0 | 0.0 |
| Chinese | Danish | eGeMAPS | SVM | 0.34 | 0.51 | 0.51 | 0.0 | 1.0 | 0.0 |
| Chinese | German | Covarep | Catboost | 0.42 | 0.42 | 0.4 | 0.43 | 0.33 | 0.51 |
| Chinese | German | Covarep | SVM | 0.46 | 0.51 | 0.53 | 0.51 | 0.2 | 0.83 |
| Chinese | German | eGeMAPS | Catboost | 0.33 | 0.5 | 0.5 | 0.0 | 1.0 | 0.0 |
| Chinese | German | eGeMAPS | SVM | 0.33 | 0.5 | 0.5 | 0.0 | 0.99 | 0.0 |
| Danish | Chinese | Covarep | Catboost | 0.41 | 0.52 | 0.51 | 0.69 | 0.96 | 0.09 |
| Danish | Chinese | Covarep | SVM | 0.51 | 0.51 | 0.51 | 0.51 | 0.55 | 0.48 |
| Danish | Chinese | eGeMAPS | Catboost | 0.4 | 0.46 | 0.47 | 0.39 | 0.75 | 0.16 |
| Danish | Chinese | eGeMAPS | SVM | 0.39 | 0.51 | 0.6 | 0.51 | 0.07 | 0.95 |
| Danish | German | Covarep | Catboost | 0.42 | 0.45 | 0.47 | 0.42 | 0.68 | 0.23 |
| Danish | German | Covarep | SVM | 0.41 | 0.53 | 0.52 | 0.92 | 0.99 | 0.07 |
| Danish | German | eGeMAPS | Catboost | 0.6 | 0.62 | 0.72 | 0.58 | 0.39 | 0.85 |
| Danish | German | eGeMAPS | SVM | 0.57 | 0.6 | 0.76 | 0.57 | 0.31 | 0.9 |
| German | Chinese | Covarep | Catboost | 0.5 | 0.5 | 0.5 | 0.5 | 0.54 | 0.47 |
| German | Chinese | Covarep | SVM | 0.57 | 0.58 | 0.56 | 0.6 | 0.69 | 0.47 |
| German | Chinese | eGeMAPS | Catboost | 0.57 | 0.59 | 0.65 | 0.57 | 0.4 | 0.79 |
| German | Chinese | eGeMAPS | SVM | 0.37 | 0.51 | 0.62 | 0.5 | 0.04 | 0.98 |
| German | Danish | Covarep | Catboost | 0.62 | 0.62 | 0.68 | 0.59 | 0.51 | 0.75 |
| German | Danish | Covarep | SVM | 0.41 | 0.48 | 0.49 | 0.48 | 0.12 | 0.86 |
| German | Danish | eGeMAPS | Catboost | 0.59 | 0.6 | 0.59 | 0.61 | 0.71 | 0.48 |
| German | Danish | eGeMAPS | SVM | 0.44 | 0.52 | 0.62 | 0.5 | 0.15 | 0.91 |

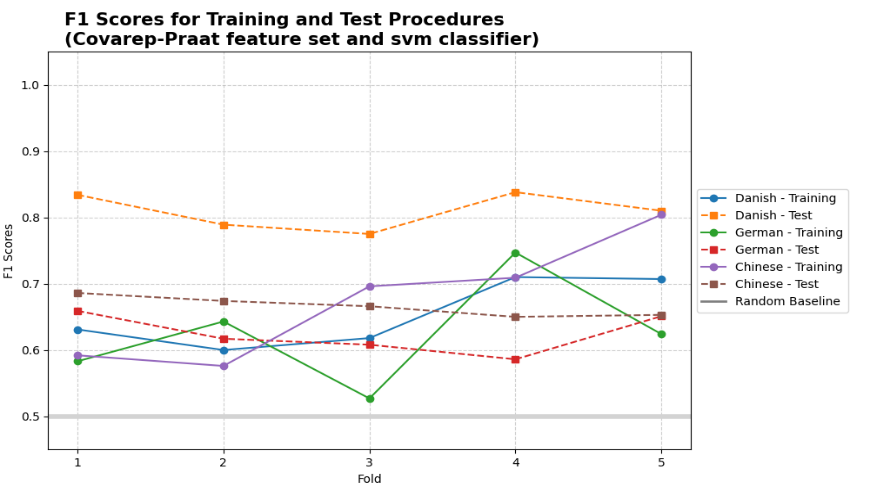

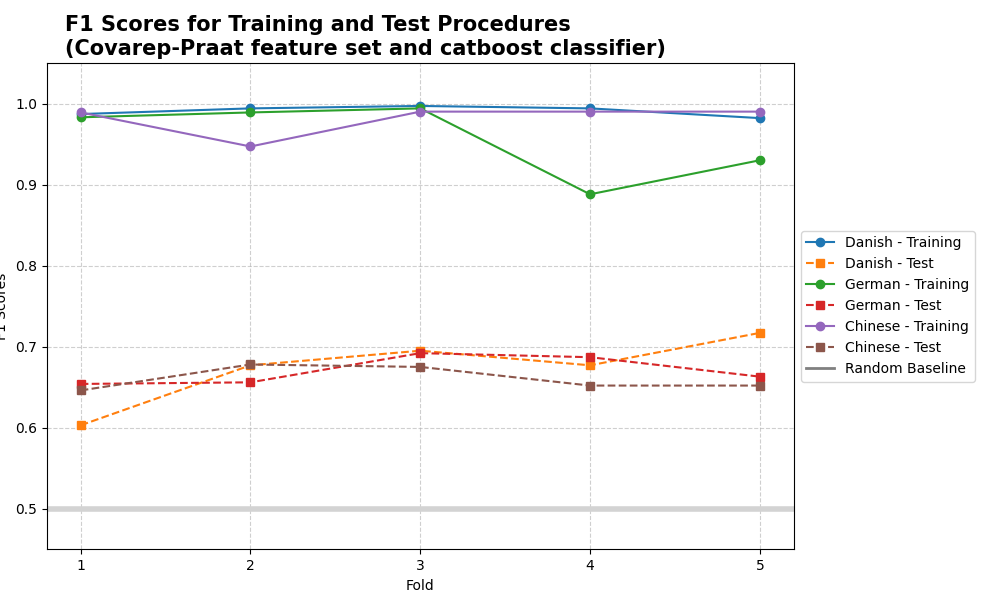

**Figure SM5_A**. Test performance (F1 score) in the training and test set for each of the 5 models trained in the 5-folds cross-validation procedure, for the Covarep features set. On the left plot is represented the performance of Catboost classifier, and on the right plot the SVM classifier.

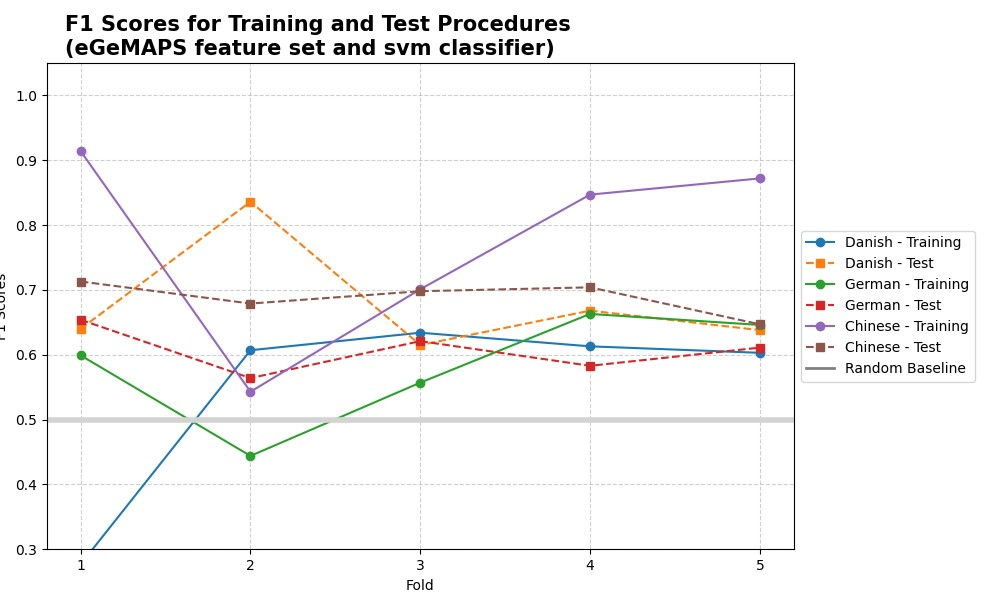

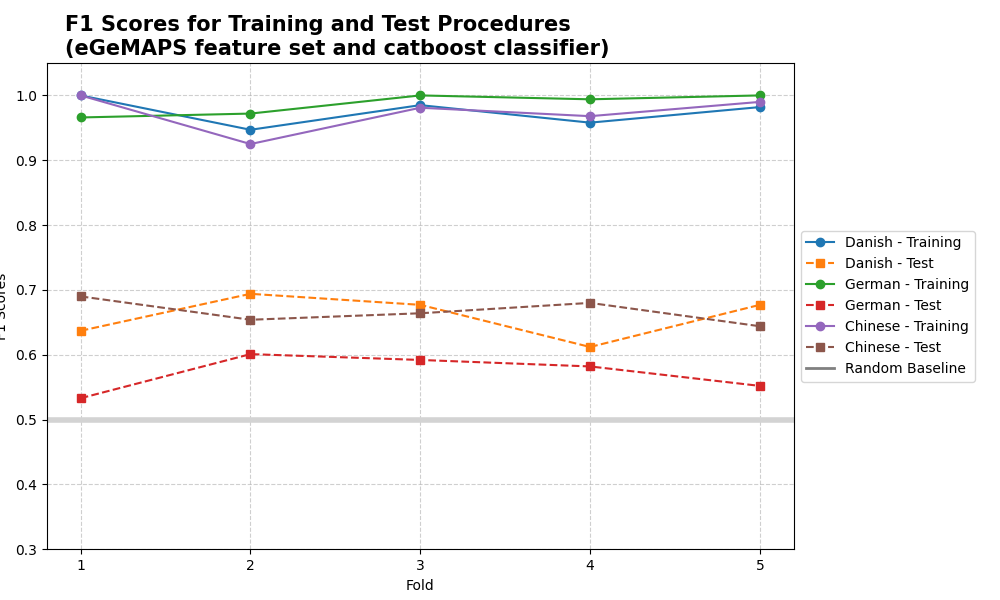

**Figure SM5_B**. Test performance (F1 score) in the training and test set for each of the 5 models trained in the 5-folds cross-validation procedure, for the eGeMAPS features set. The left plot represents the performance of the Catboost classifier, and the right plot the SVM classifier.

**MoE models**

We provide below a series of plots for each Mixture of experts (MoE) models. MoE models represent the weighted mixture of the predictions produced by the experts that were trained in different monolingual corpora. On the top line of each plot, it is represented the performance of each monolingual model, i.e. expert, in predicting the new participants (see section “Model training” for details on how the models were trained) of a different corpus speaking a different language. For example, in the Figure SM5_C, the top left plot represents the model trained on Danish participants predicting new participants speaking German, and the top right plot the model trained on Chinese participants predicting new participants speaking German. The two bar plots show how many new unseen participants have been classified as either patients or controls by the two expert models, and the relative confidence (mean +- sd). On the bottom plot, you can see the predictions by the MoE model, which is the weighted mixture of the predictions produced by the two experts above. For example, in the figure SM5_C, the MoE model (bottom line) is predicting new participants as mostly patients with schizophrenia, since the expert on the top right trained on Chinese participants is overconfident that most of the new participants are patients, and it thus biased the predictions of the MoE model toward the “patients” category.

***Figure SM5_C. MoE and monolingual models predicting new participants speaking German. The models employed a Catboost classifier trained on the eGeMAPS feature set.***

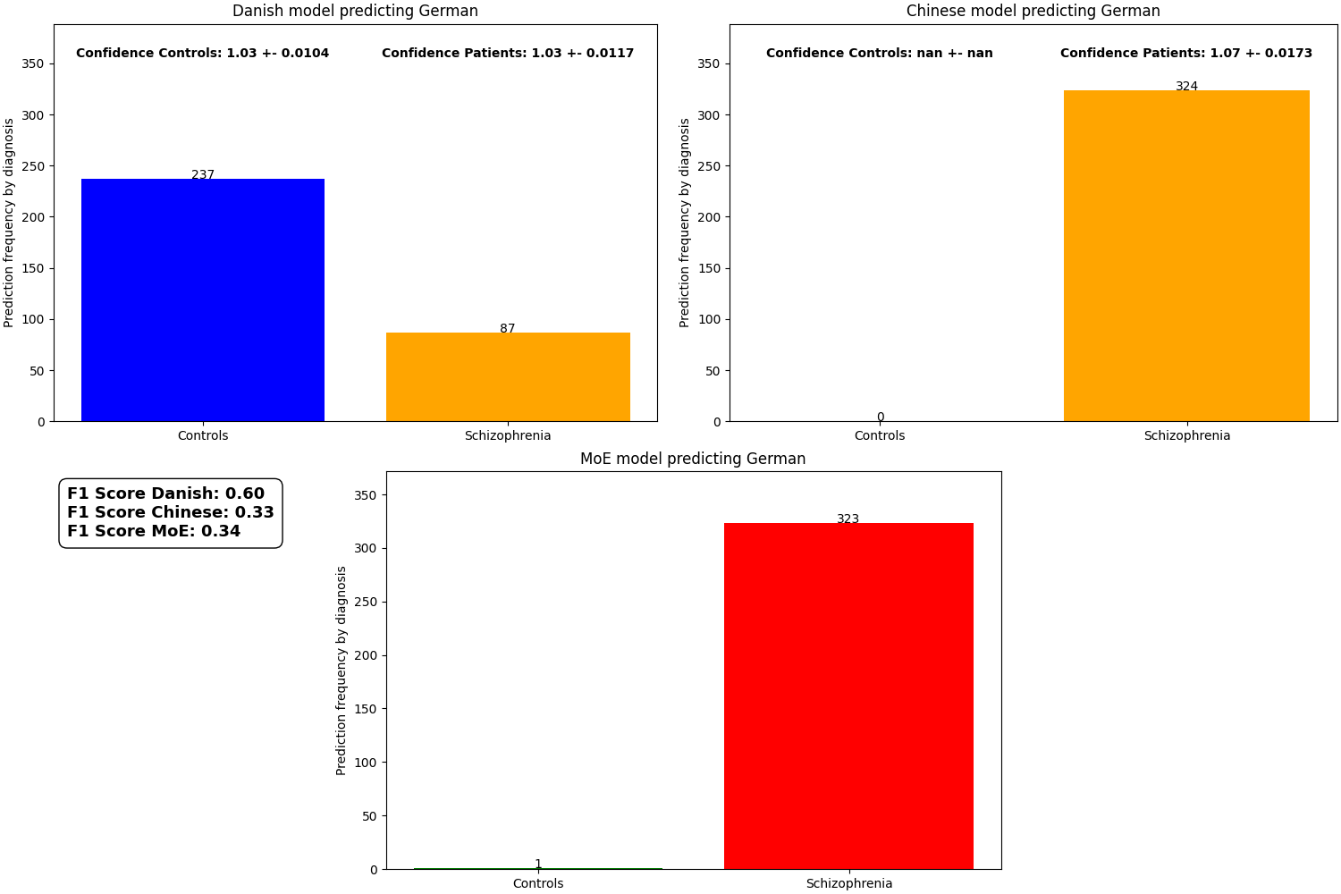

**Figure SM5_D. MoE and monolingual models predicting new participants speaking German. The models used a SVM classifier trained on the eGeMAPS feature set.**

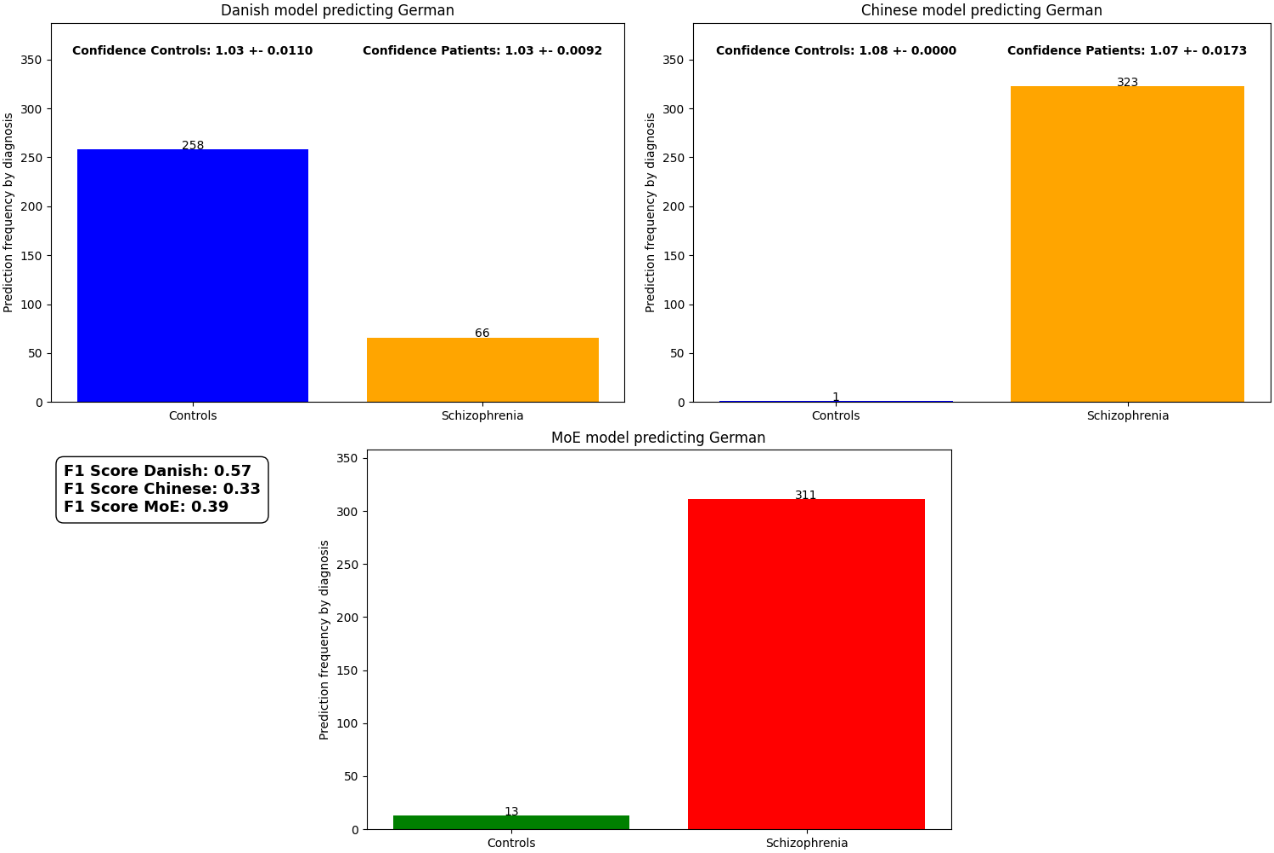

***Figure SM5_E. MoE and monolingual models predicting new participants speaking German. The models were trained with Catboost classifier and Covarep feature set.***

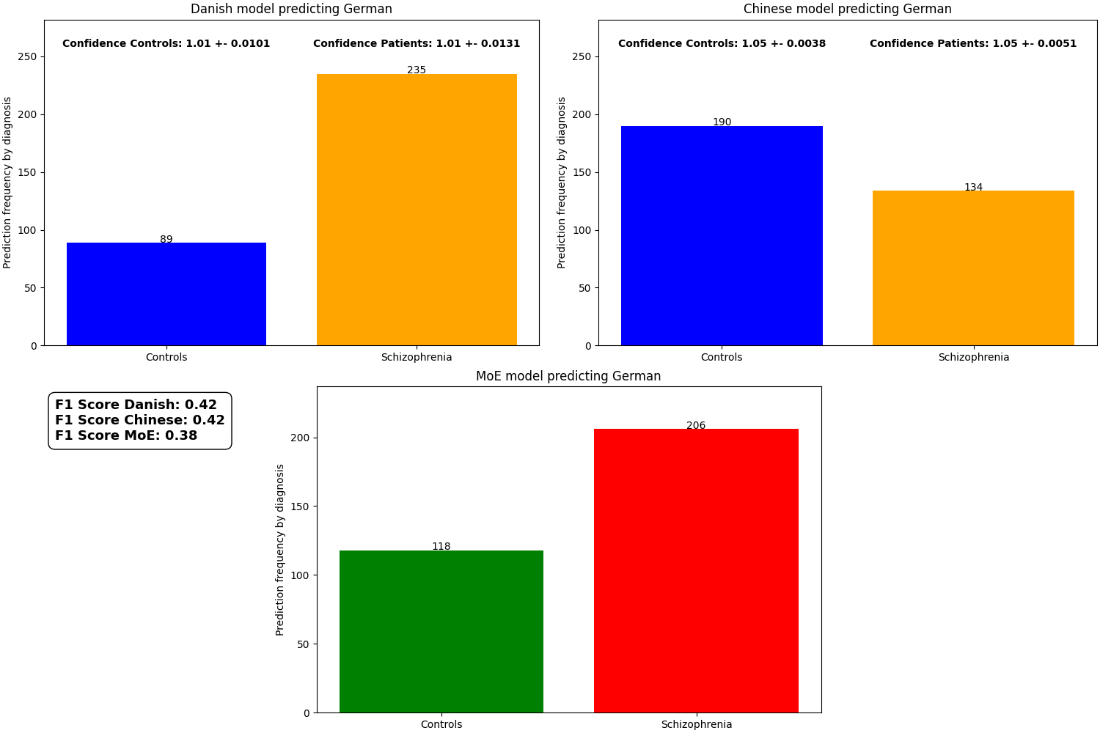

***Figure SM5_F. MoE and monolingual models predicting new participants speaking German. The models were trained with SVM classifier and Covarep feature set.***

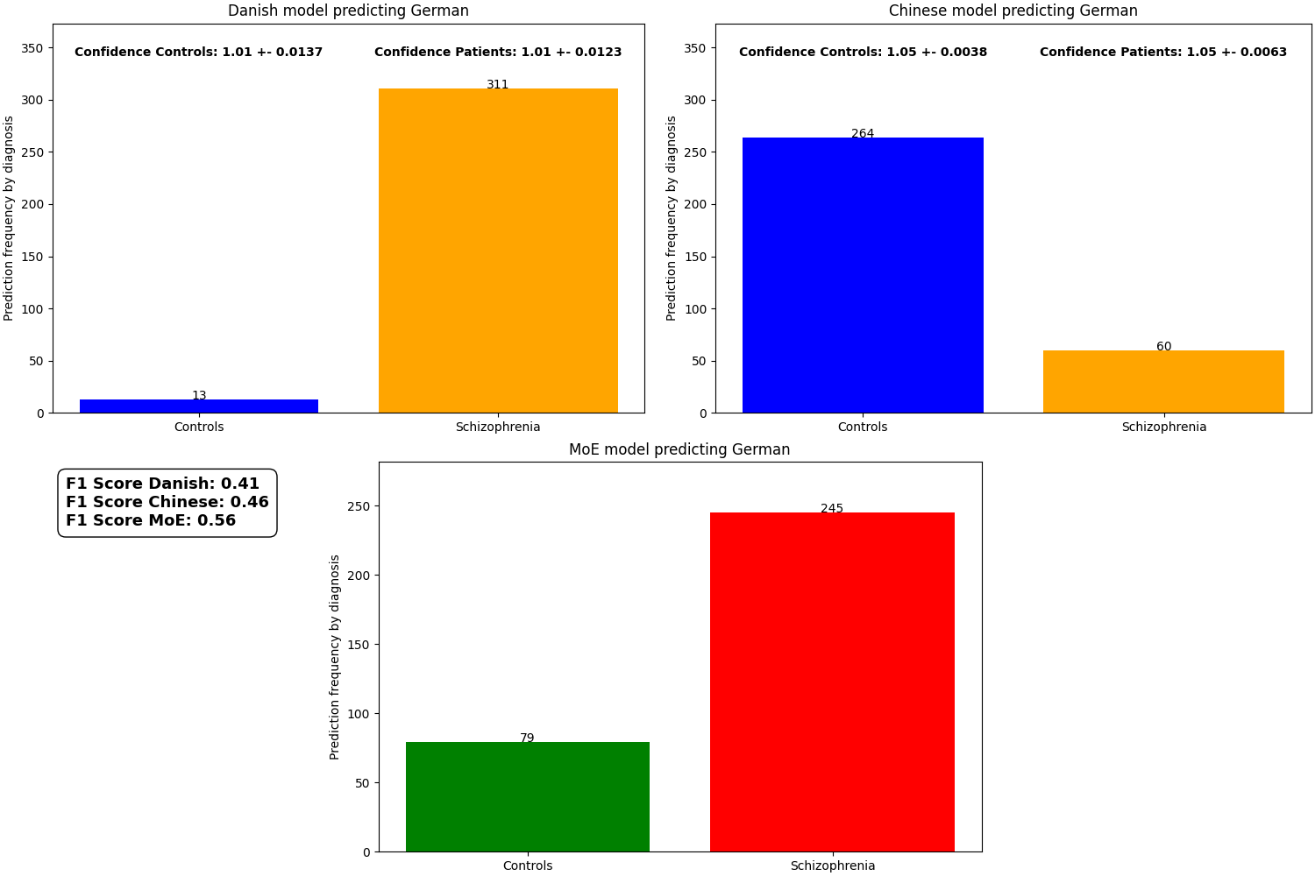

***Figure SM5_G. MoE and monolingual models predicting new participants speaking Chinese. The models were trained with Catboost classifier and eGeMAPS feature set.***

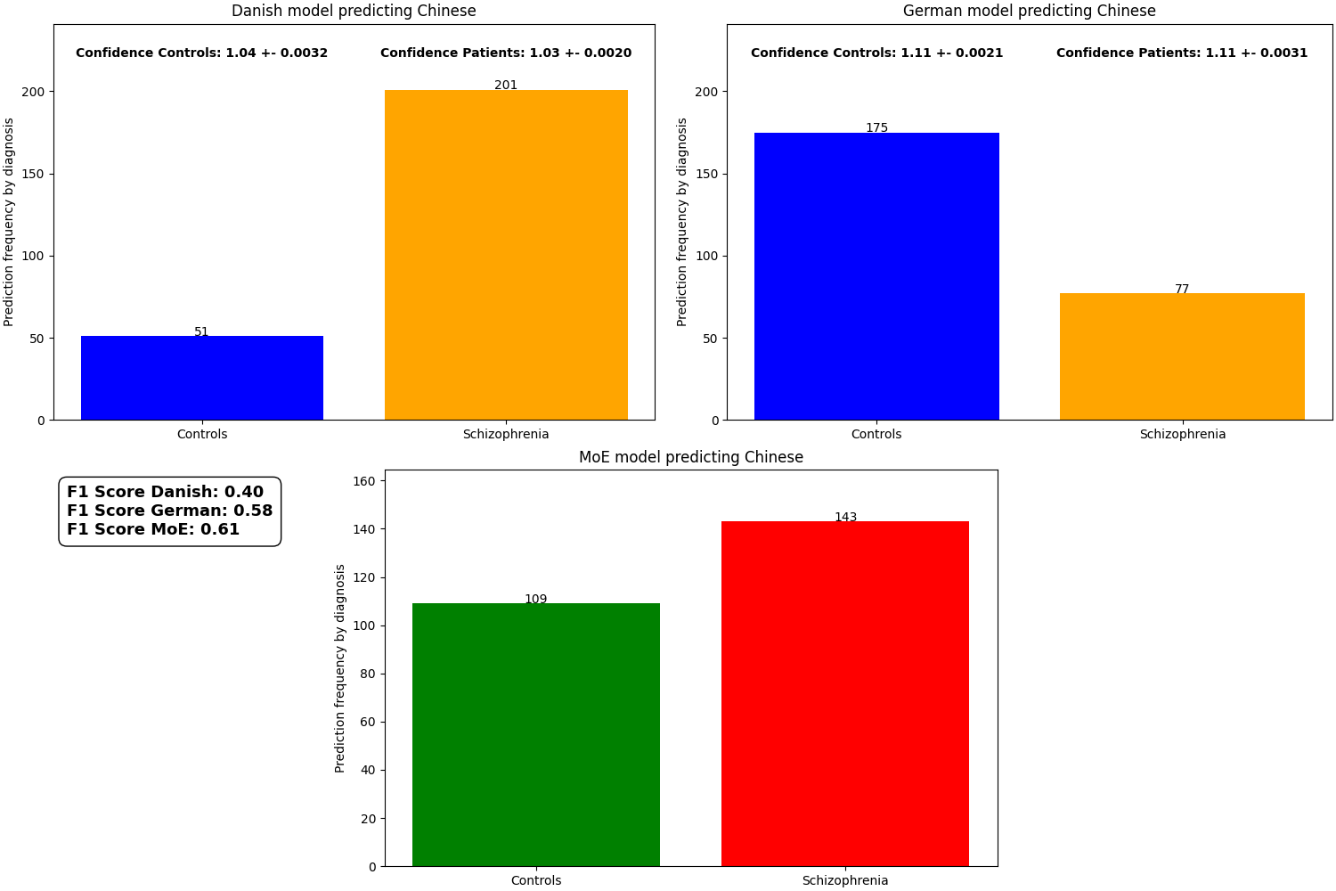

**Figure SM5_H. MoE and monolingual models predicting new participants speaking Chinese. The models were trained with SVM classifier and eGeMAPS feature set.**

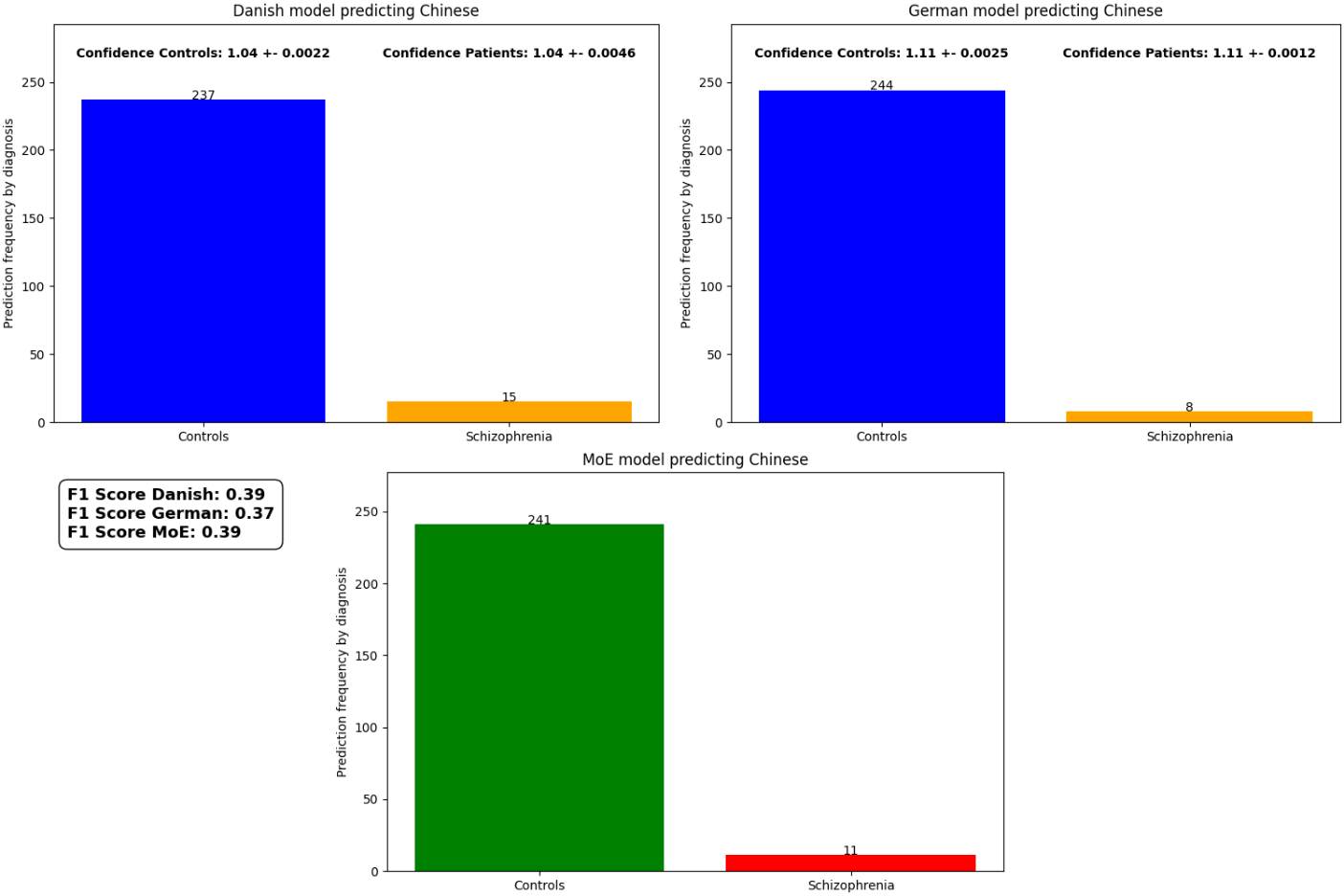

***Figure SM5_I. MoE and monolingual models predicting new participants speaking Chinese. The models were trained with Catboost classifier and Covarep feature set.***

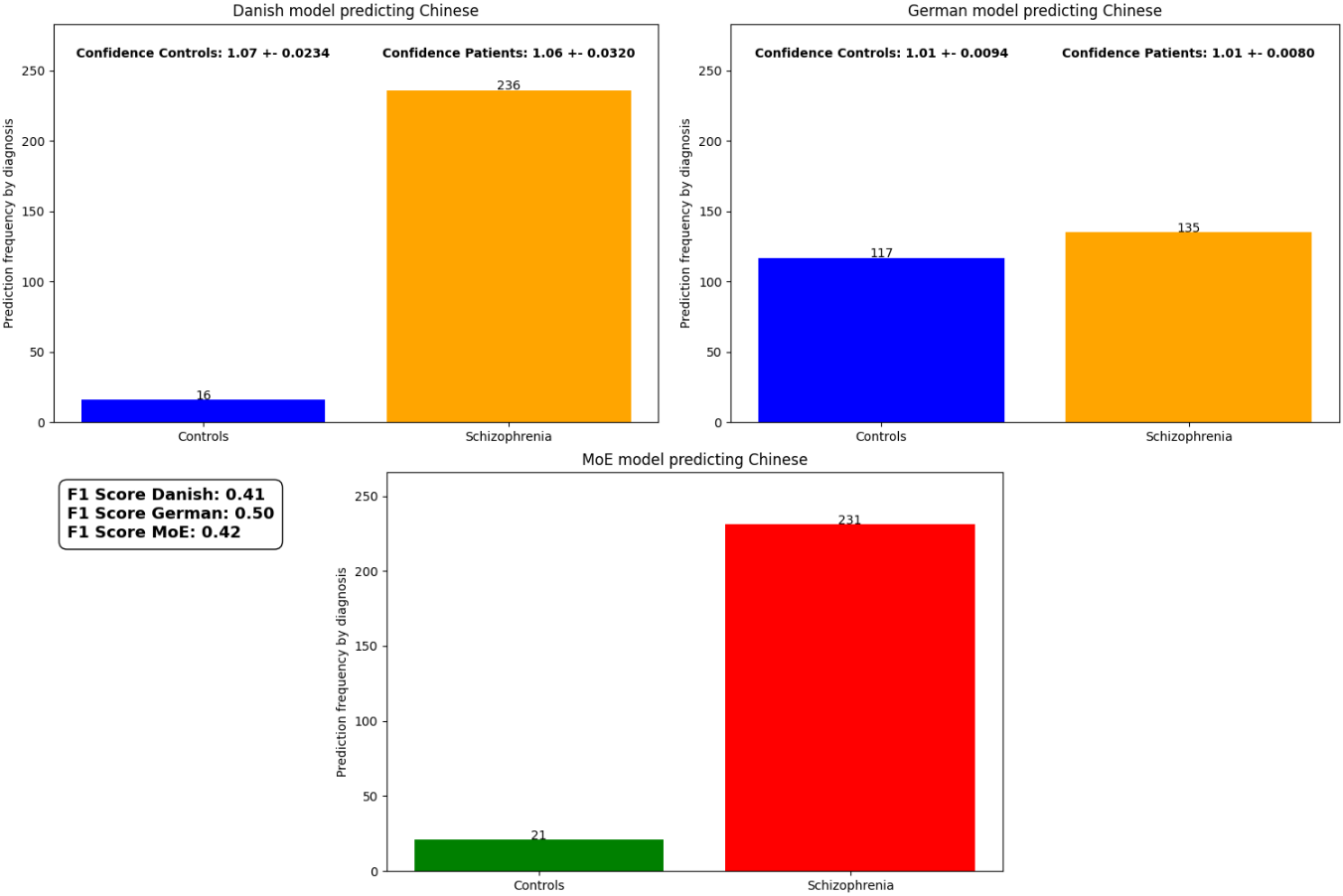

***Figure SM5_L. MoE and monolingual models predicting new participants speaking Chinese. The models were trained with SVM classifier and Covarep feature set.***

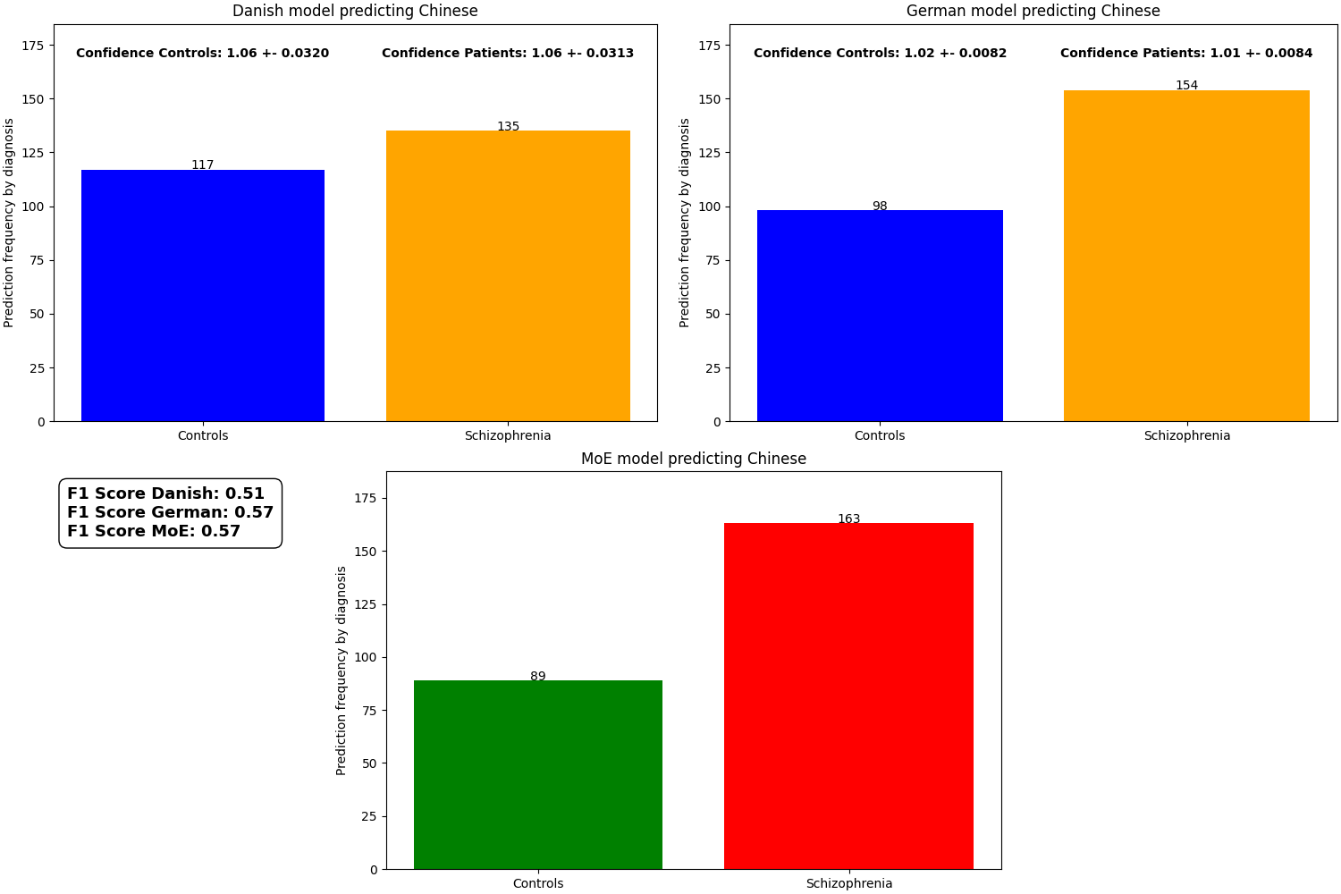

***Figure SM5_M. MoE and monolingual models predicting new participants speaking Danish. The models were trained with Catboost classifier and eGeMAPS feature set.***

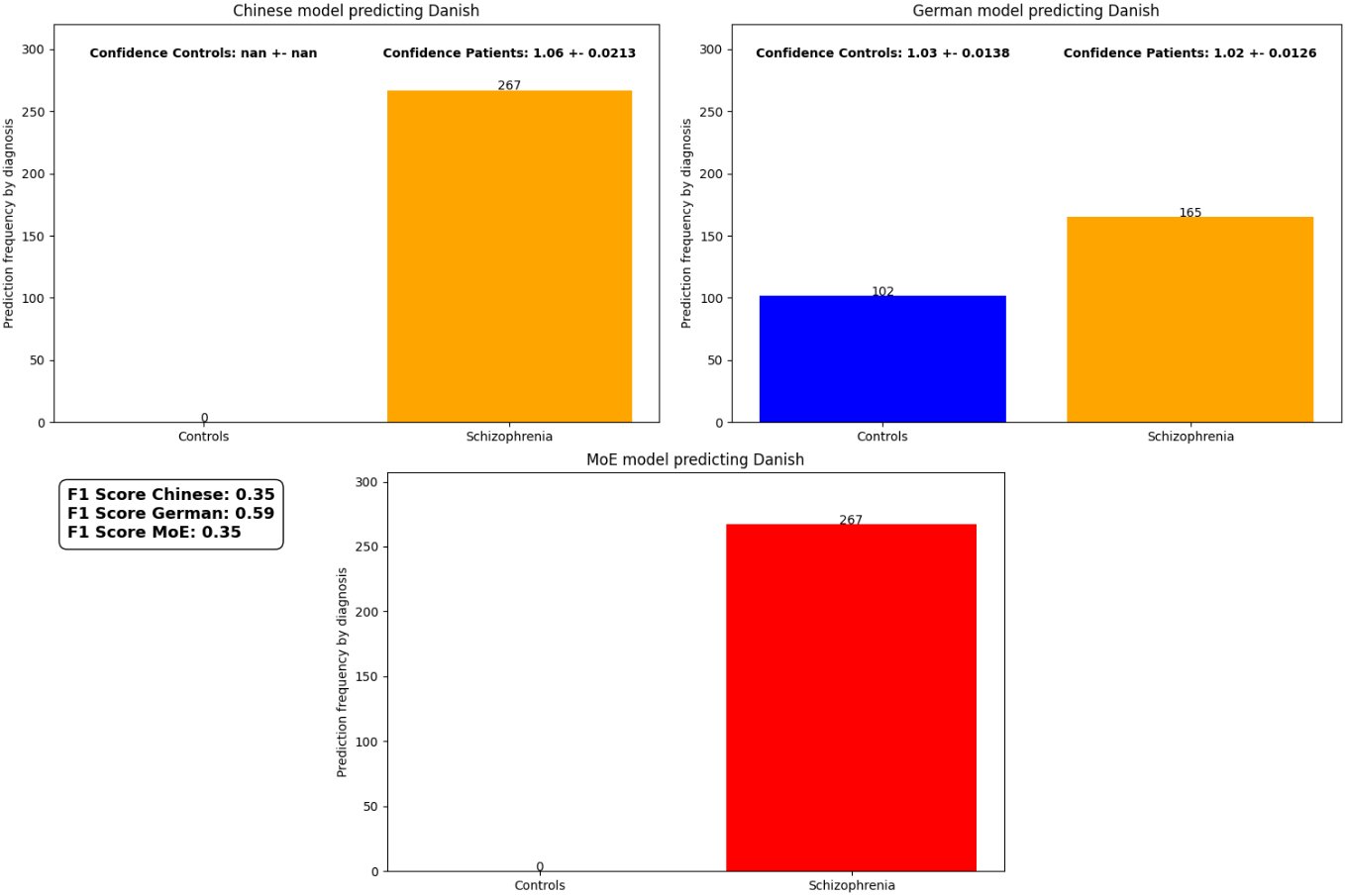

***Figure SM5_N. MoE and monolingual models predicting new participants speaking Danish. The models were trained with SVM classifier and eGeMAPS feature set.***

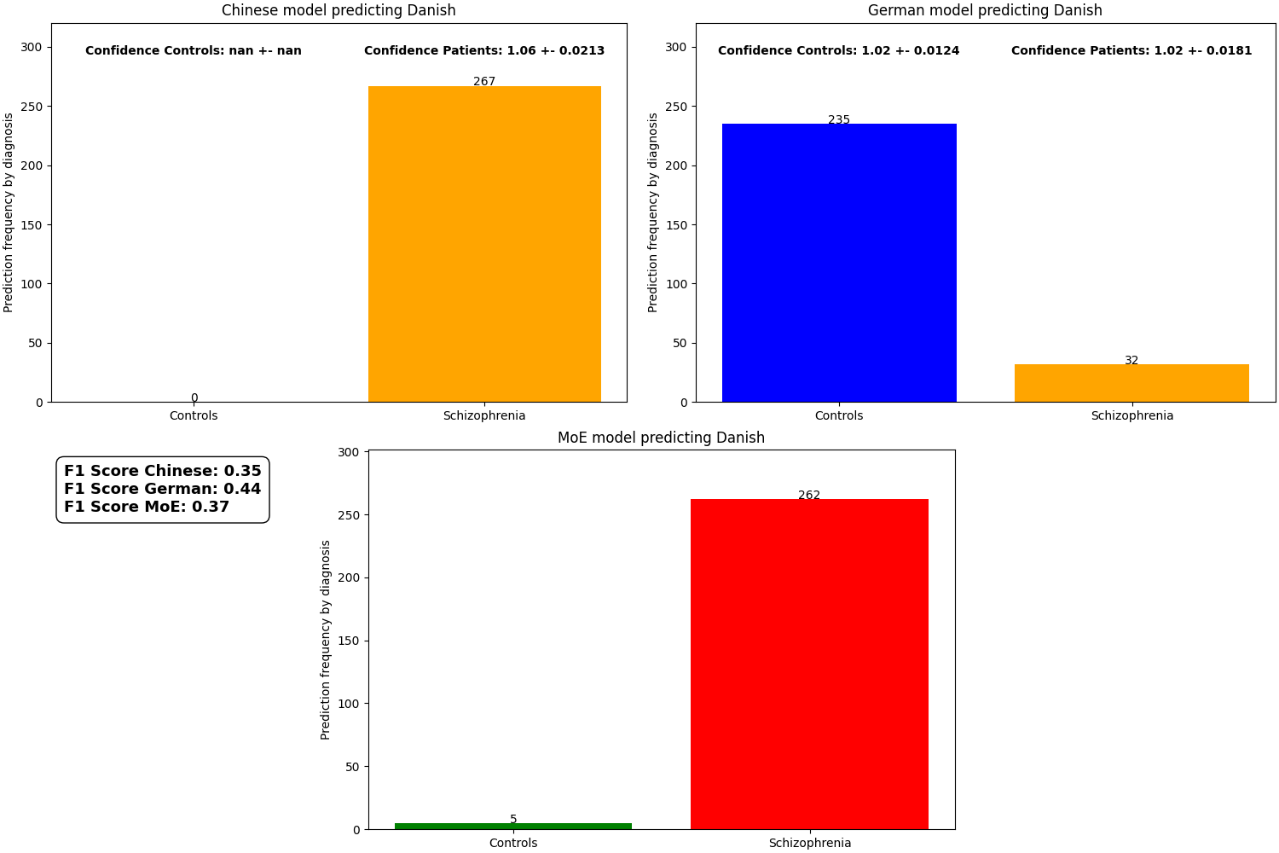

***Figure SM5_O. MoE and monolingual models predicting new participants speaking Danish. The models were trained with Catboost classifier and Covarep feature set.***

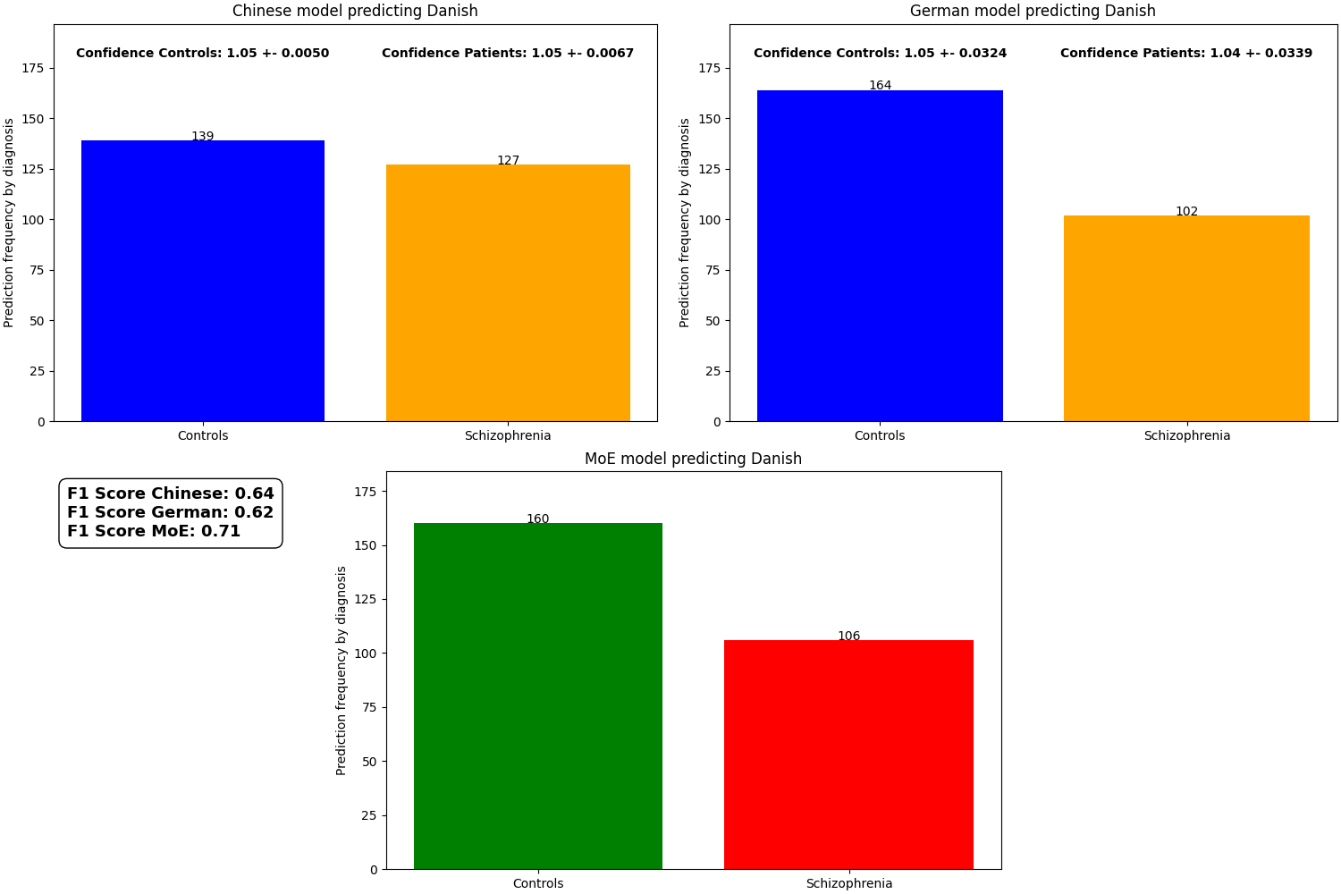

***Figure SM5_P. MoE and monolingual models predicting new participants speaking Danish. The models were trained with SVM classifier and Covarep feature set.***

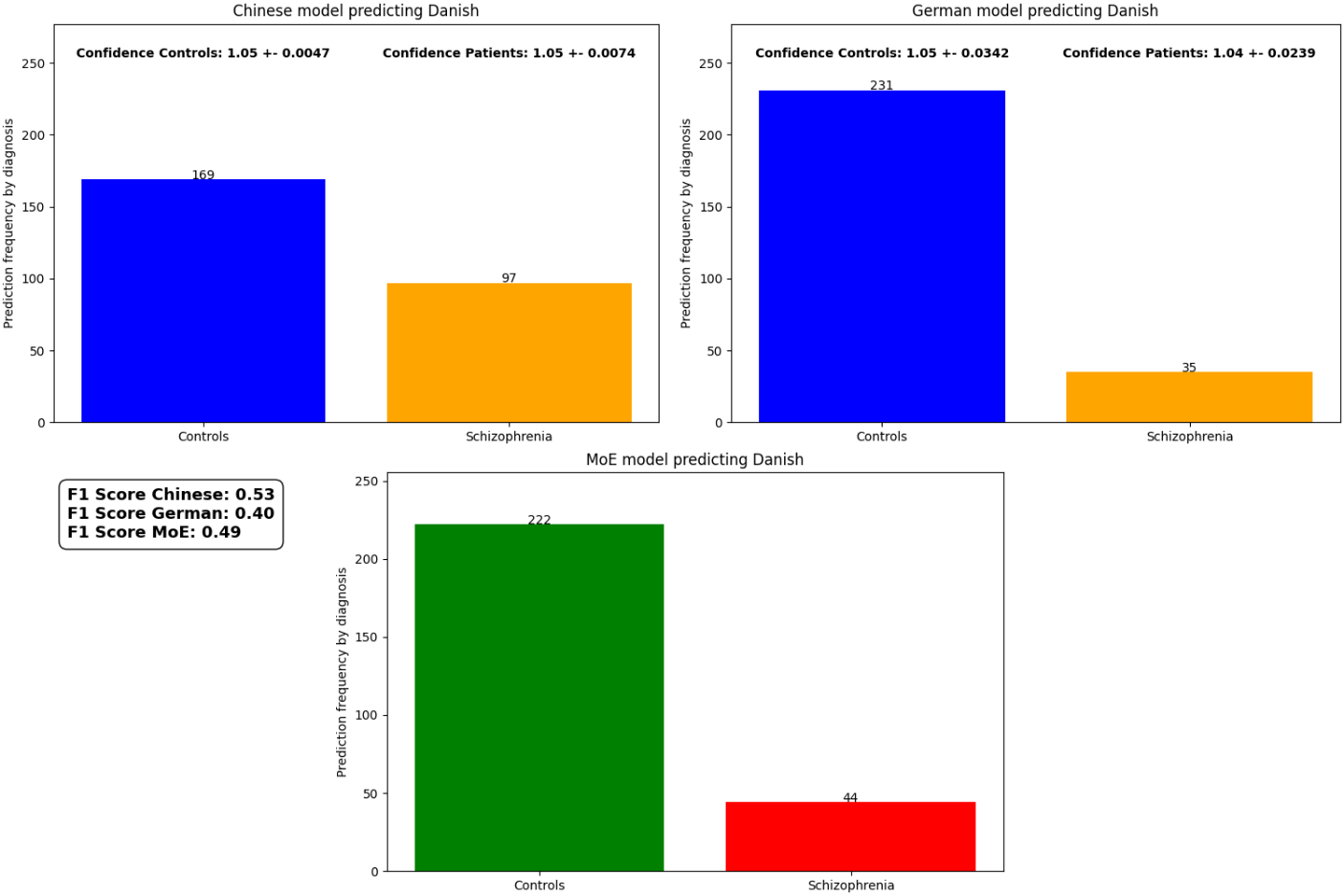

**SM6 - Features importance**

In this section we provide supplementary analysis to assess the most important features used by each model for predicting the diagnosis of new participants speaking the same (Q1) or a different language (Q2). To assess feature importance, we used the permutation feature importance method which works by randomly shuffling each feature and measuring the change in the model’s performance. A feature is considered important if shuffling its values decreases the model performance because the model relies on this feature for making predictions.

**Q1:** Overall, we have observed a low consistency and high variability in which acoustic features were most relevant for the different models trained and tested on the same language. For example, while F0 related features (F0 standard deviation, F0 semitones) showed to be important for classifying new participants speaking German see Fig. S5_C), they were not among the most important features in the other two models (Danish and Chinese, see Fig. S5_A and Fig. S5_B). Or while jitter and shimmer were important for classifying new participants speaking Danish (see Fig Fig. S5_A), they were not important in the models predicting new Chinese participants (see Fig. Fig. S5_B).

Further, the model trained with different classifiers (Catboost and SVM) and on different feature sets (eGeMAPS and Covarep) showed only a partial consistency between the most relevant features. For example, the “voiced segment per seconds” feature was among the most important features for the model classifying new participants speaking Danish with the Catboost classifier and the eGeMAPS features set, but was not important for the Danish model with the SVM classifier and the eGeMAPS features (see Fig. S5_A).

Overall, this might suggests that the models might not be capturing diagnosis- or symptoms-specific markers of schizophrenia, but rather learning heuristics that may only be effective for samples with comparable socio-demographic and clinical features, or under similar recording conditions.

***Figure S5_A. Models trained and tested on participants speaking Danish.***

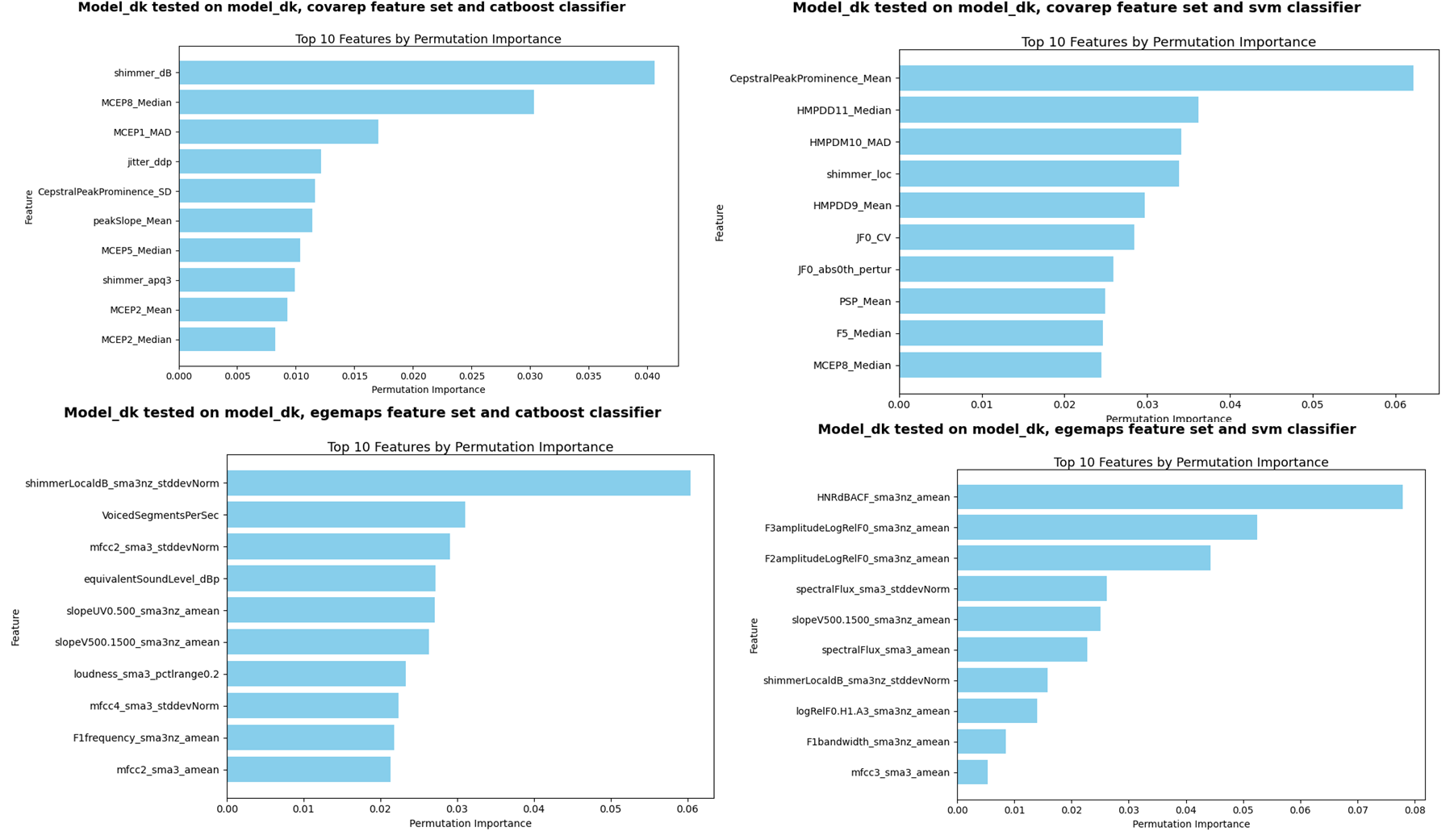

***Figure S5_B. Models trained and tested on participants speaking Chinese.***

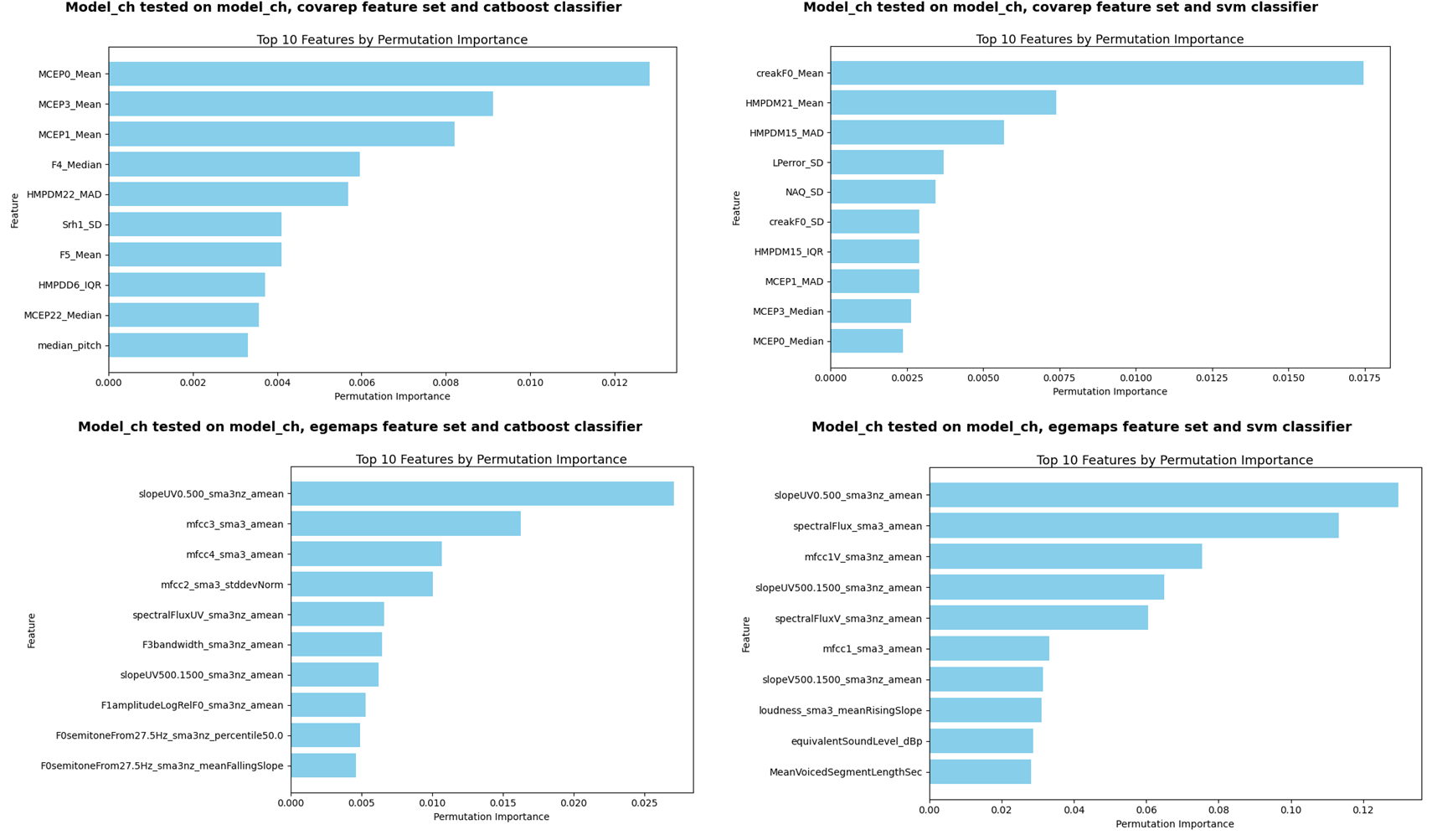

***Figure S5_C. Models trained and tested on participants speaking German.***

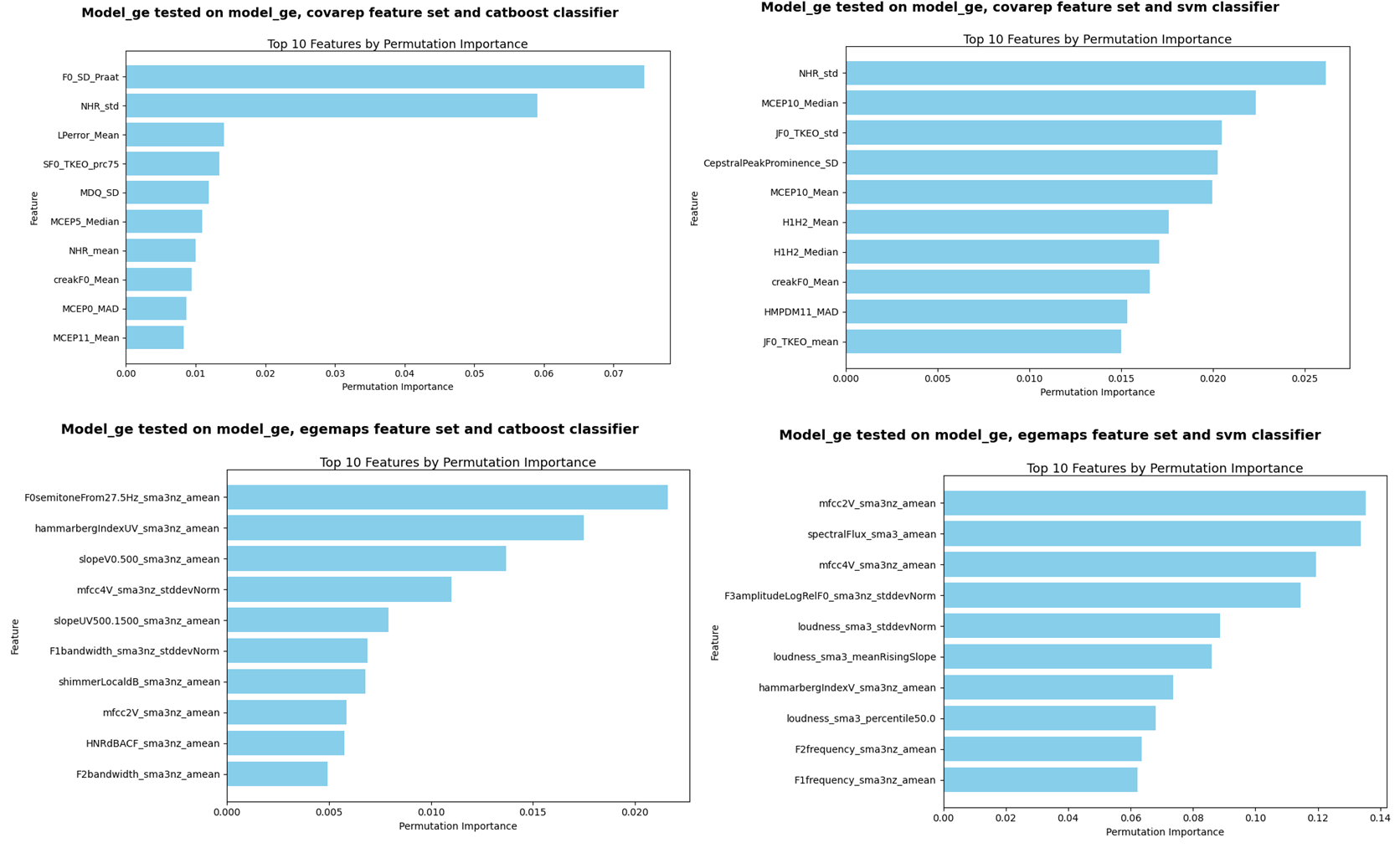

**Q2:** Overall, and in line with the pattern observed for the models predicting participants speaking the same language (Q1), we observed a low consistency and high variability in which acoustic features were most relevant for the different models trained and tested on different languages.

***Figure S5_D. Models trained on participants speaking German and tested on participants speaking Danish.***

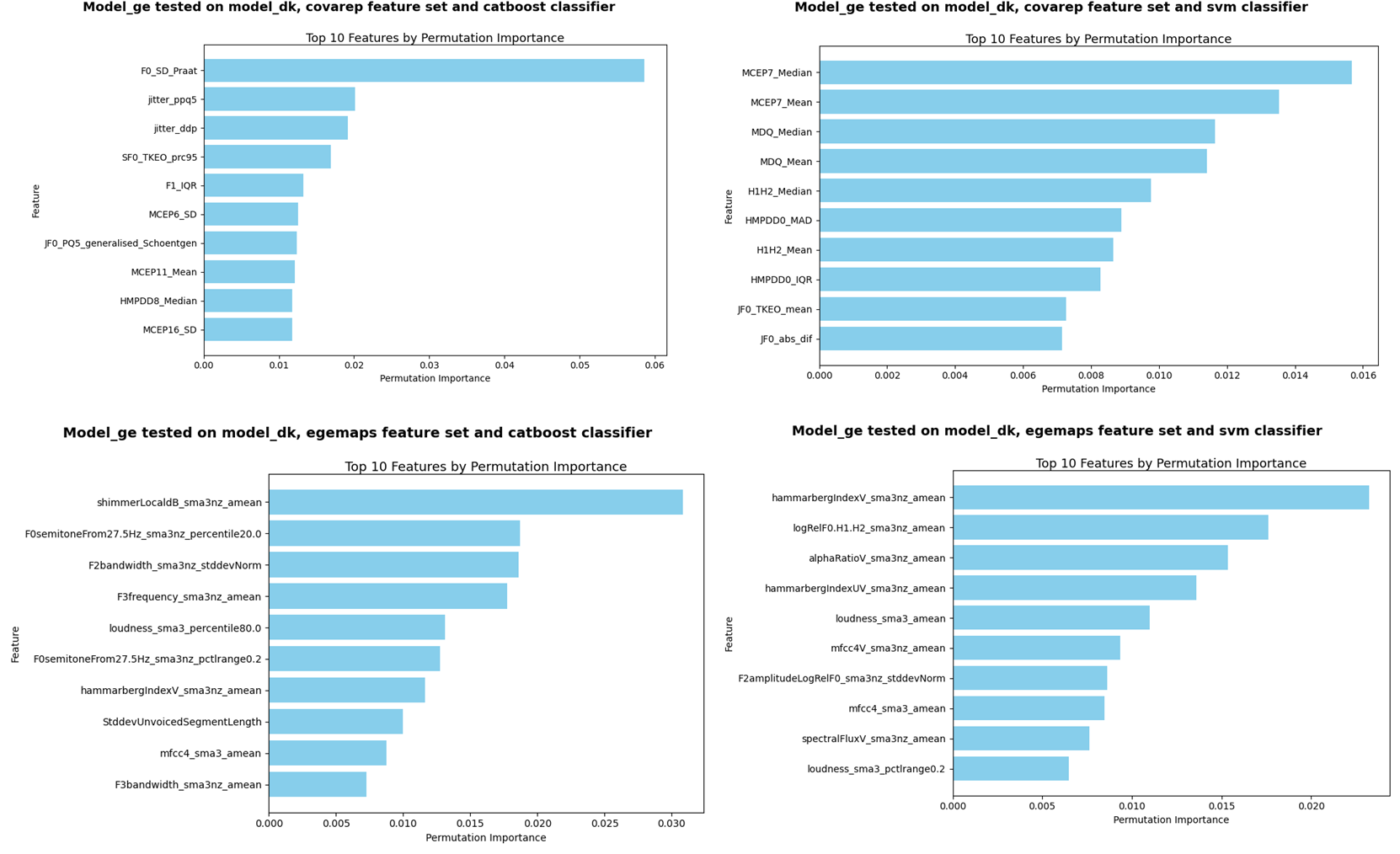

***Figure S5_E. Models trained on participants speaking German and tested on participants speaking Chinese.***

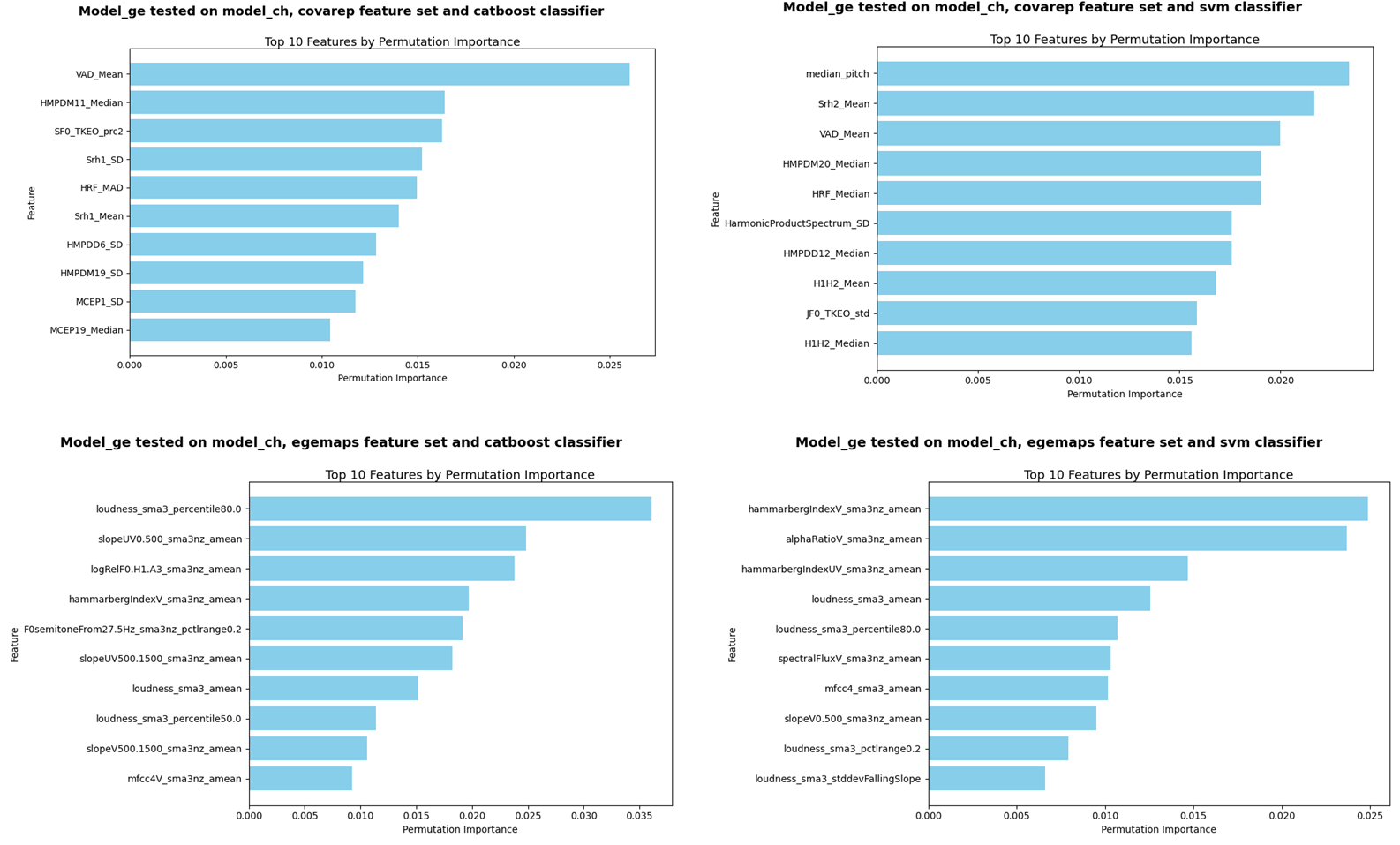

***Figure S5_F. Models trained on participants speaking Danish and tested on participants speaking Chinese.***

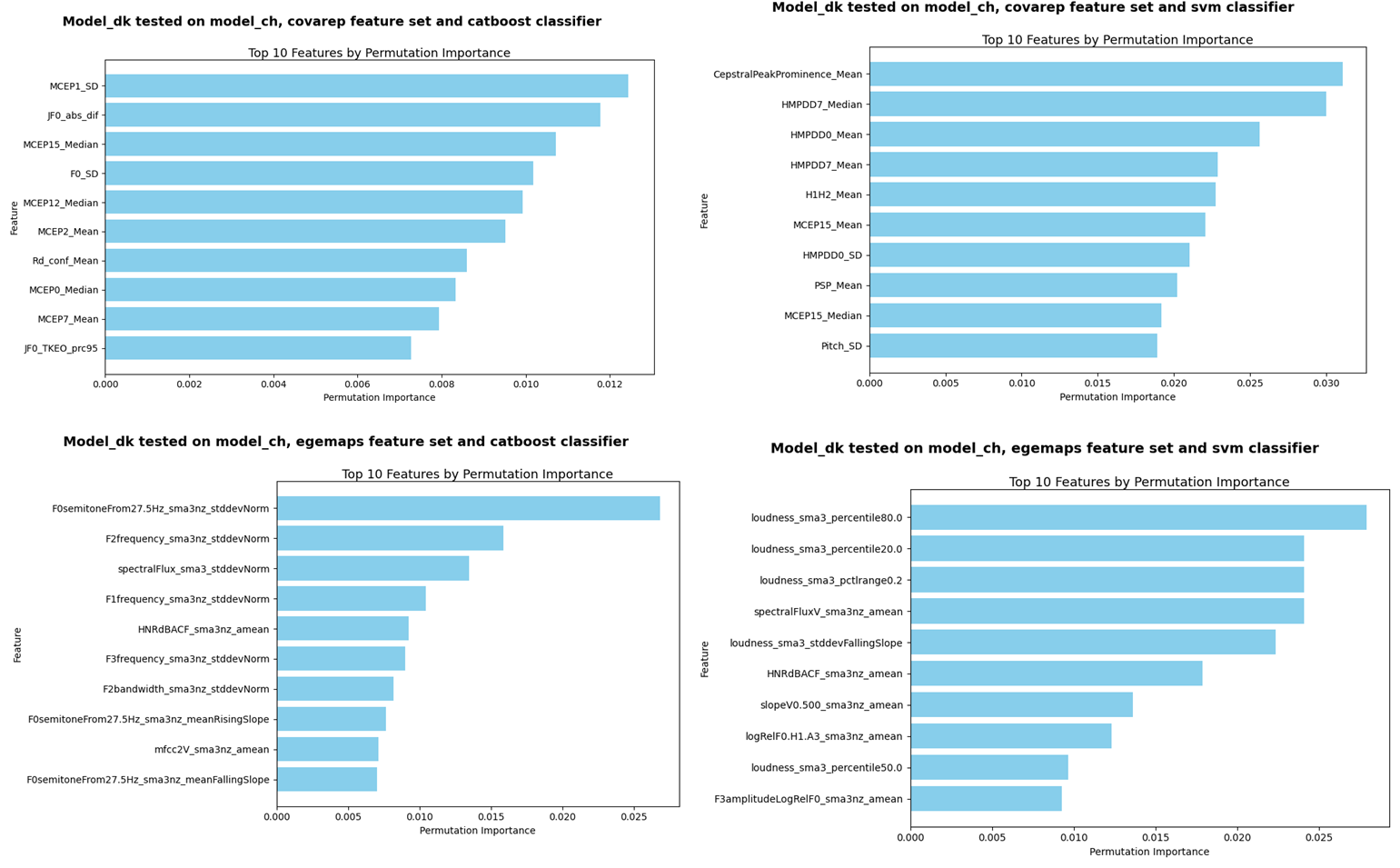

***Figure S5_G. Models trained on participants speaking Danish and tested on participants speaking German.***

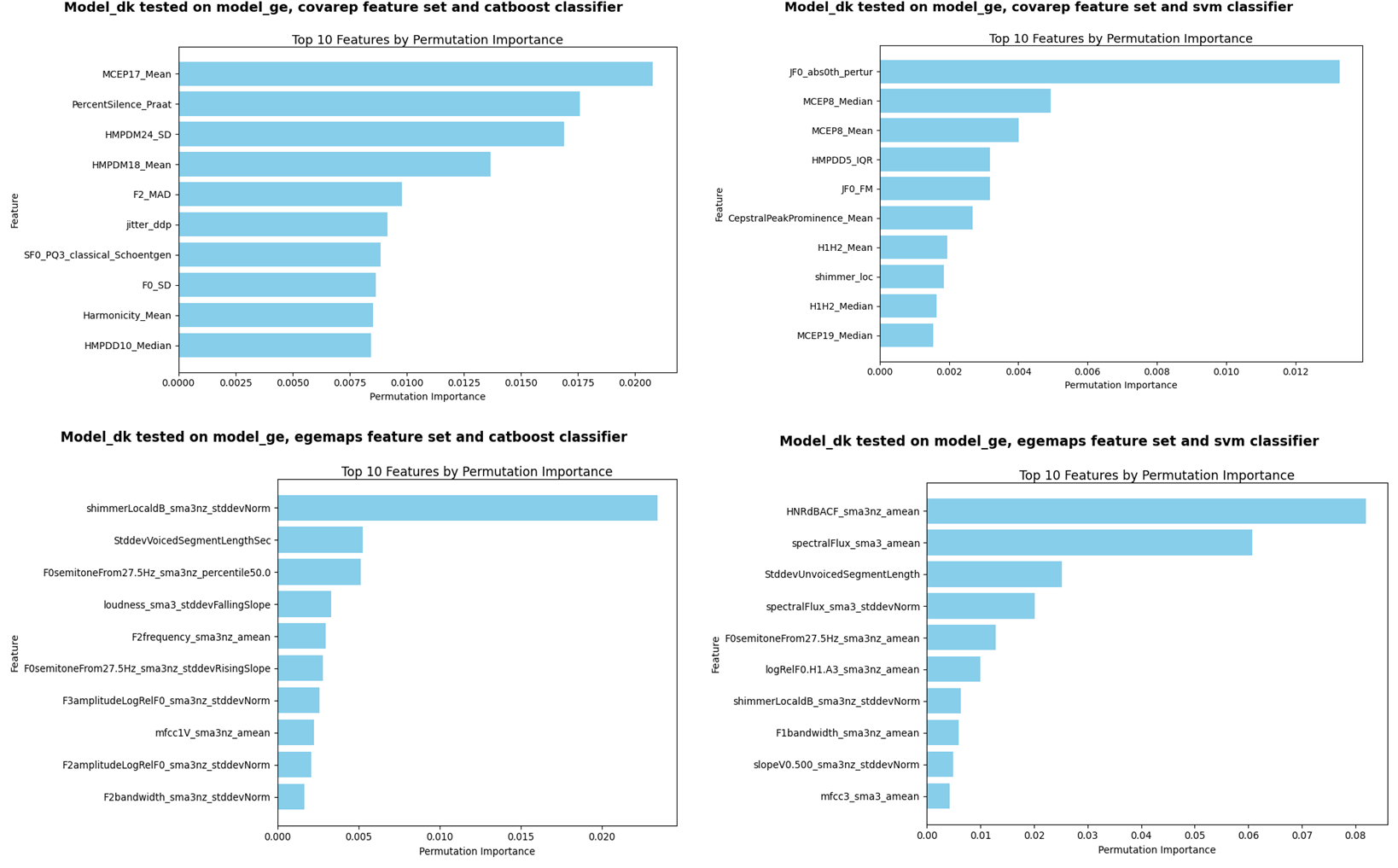

***Figure S5_H. Models trained on participants speaking Chinese and tested on participants speaking Danish.***

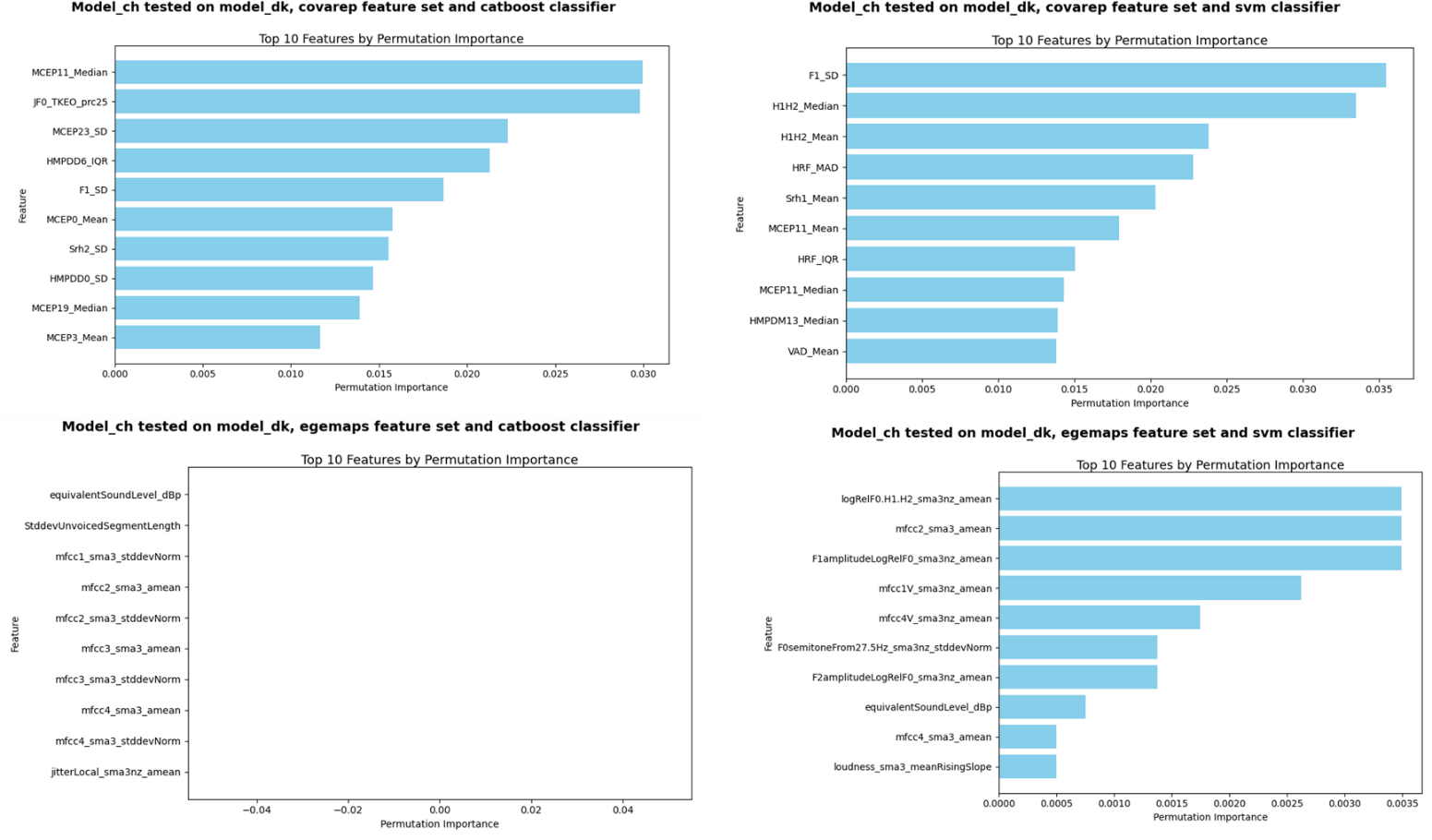

***Figure S5_I. Models trained on participants speaking Chinese and tested on participants speaking German.***

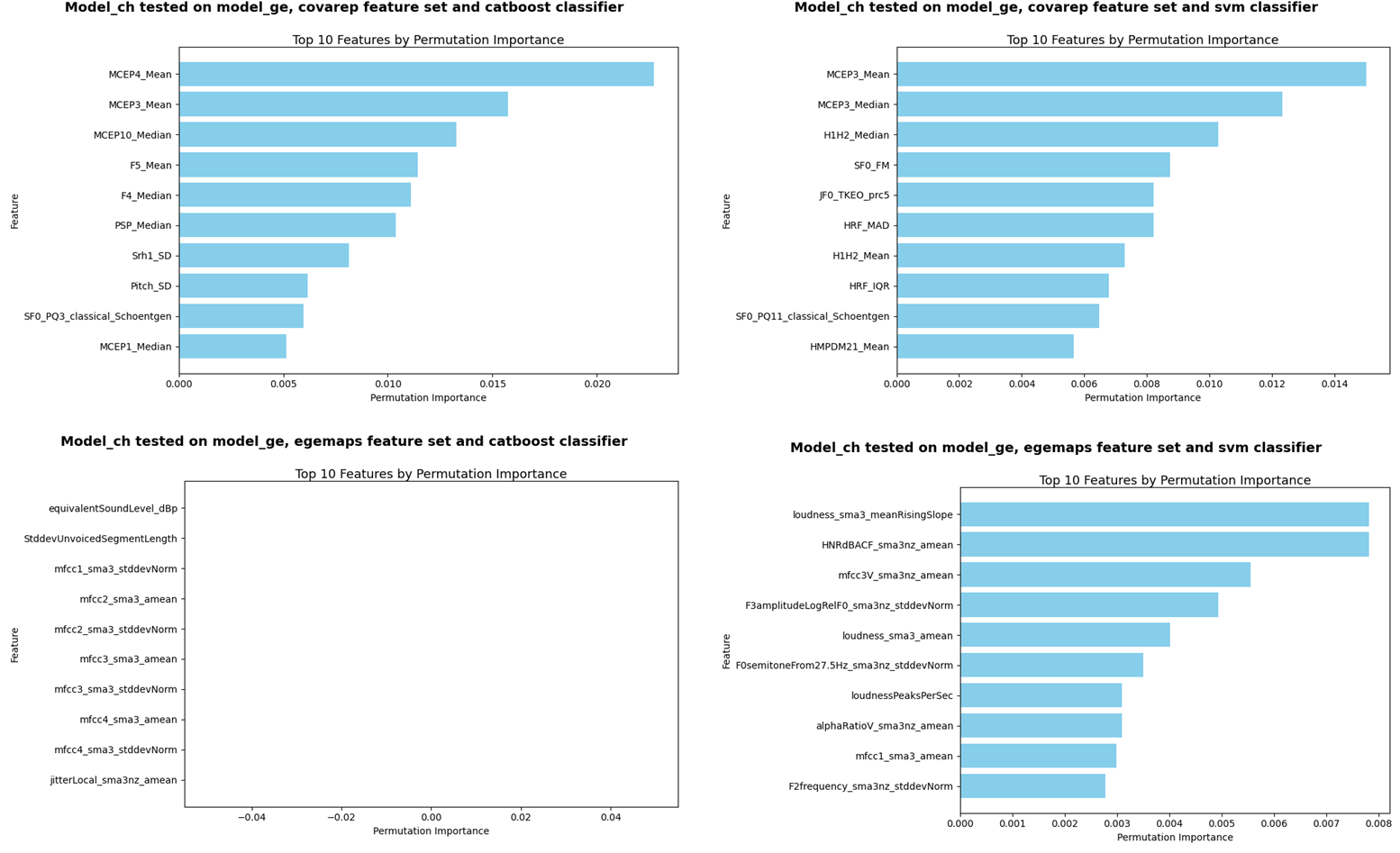

**SM7 – Comparison of models trained and tested on similar or different language families**

In this section we provide additional analysis comparing the performance of models trained and tested on participants speaking languages of the same language family (e.g. Danish and German), compared to models trained and tested on participants speaking languages of different language families (e.g., Danish and Chinese).

We have not observed a clear improvement in generalization performance of models trained and tested on participants speaking languages of the same language family (e.g. Danish and German), compared to models trained and tested on participants speaking languages of different language families (e.g., Danish and Chinese). As can be seen in Fig S6_A, while the models trained and tested on Danish or German languages showed a small improvement compared to models trained on Danish or German and tested on Chinese, or trained on Chinese and tested on Danish or German, their performance is still at chance level.

***Figure SM7_A.*** *Performance score (F1) of models trained and tested on participants speaking languages of the same language family, i.e. Germanic, compared to models trained and tested on participants speaking languages of different language families, i.e. not Germanic.*

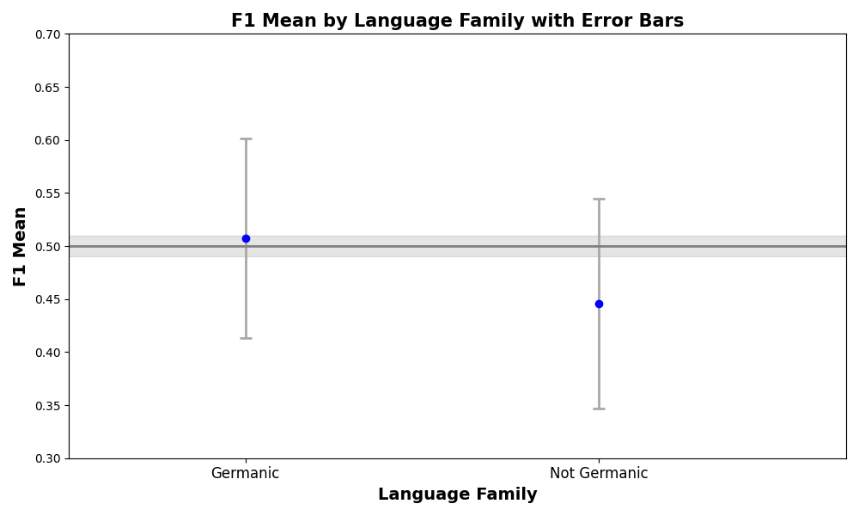
